# A new robust and accurate two-sample Mendelian randomization method with a large number of genetic variants

**DOI:** 10.1101/2024.04.16.24305933

**Authors:** Lei Zhang, Jun-Jie Niu, Xian-Mei He, Xiao Zheng, Qi-Gang Zhao, Xiu-Juan Yu, Li Luo, Hai-Gang Ren, Yu-Fang Pei

**Affiliations:** Center for Genetic Epidemiology and Genomics, School of Public Health, Suzhou Medical College of Soochow University, Jiangsu, 215123, PR China; MOE Key Laboratory of Geriatric Diseases and Immunology, Soochow University, Suzhou, Jiangsu 215123, PR China; Department of Orthopedic Surgery, the First Affiliated Hospital of Soochow University, Suzhou, Jiangsu, 215006, PR China; Department of Epidemiology and Biostatistics, School of Public Health, Suzhou Medical College of Soochow University, Suzhou, Jiangsu, 215123, PR China; School of Physical Education and Sport Science, Soochow University, Suzhou, 215021, PR China; Laboratory of Molecular Neuropathology, Jiangsu Key Laboratory of Neuropsychiatric Diseases and College of Pharmaceutical Sciences, Soochow University, Suzhou, 215123, PR China

**Keywords:** Mendelian randomization, uncorrelated pleiotropy, correlated pleiotropy, appendicular lean mass, lipid traits

## Abstract

Horizontal pleiotropy can significantly confound causal estimates in Mendelian randomization (MR) studies, particularly when numerous instrumental variables (IVs) are employed. In this study we propose a novel statistical method, Mendelian Randomization analysis based on Z-scores (MRZ), to conduct robust and accurate MR analysis in the presence of pleiotropy. MRZ models the IV-outcome association z-score as a mixture distribution, separating the causal effect of the exposure on the outcome from pleiotropic effects specific to each IV. By classifying IVs into distinct categories (valid, uncorrelated pleiotropic, and correlated pleiotropic), MRZ constructs a likelihood function to estimate both causal and pleiotropic effects. Simulation studies demonstrate MRZ’s robustness, power, and accuracy in identifying causal effects under diverse pleiotropic scenarios and overlapped samples. In a bidirectional MR analysis of appendicular lean mass (ALM) and four lipid traits using both the UK Biobank (UKB)-internal datasets and the UKB-Global Lipids Genetics Consortium (GLGC) joint datasets, MRZ consistently identified a causal effect of ALM on total cholesterol (TC) and low-density lipoprotein cholesterol (LDL-C). Conversely, existing methods often detected mutual causal relationship between lipid traits and ALM, highlighting their susceptibility to confounding by horizontal pleiotropy. A randomized controlled experiment conducted in mice validated the absence of causal effect of TC on ALM, corroborating the MRZ findings and further emphasizing its resilience against pleiotropic biases.

## Introduction

Determining whether a modifiable exposure causes a particular disease outcome is crucial for understanding disease mechanisms and guiding prevention and clinical interventions. However, traditional observational analyses often face limitations in establishing such causal effects due to potential confounding by unobserved variables or reverse causation [1]. Mendelian randomization (MR) provides a solution by offering a statistical approach to infer causation from observational data while circumventing both unobserved confounding and reverse causation [1, 2]. MR utilizes genetic variation associated with an exposure as an instrumental variable (IV) to investigate the causal effect of the exposure on an outcome [2].

MR has proven highly successful in uncovering causal relationships across a wide range of epidemiological conditions and diseases, establishing itself as a popular approach in modern genetic epidemiology [3]. The rapid growth of genome-wide association studies (GWASs) further accelerates the application of MR by providing a wealth of data. With larger sample sizes, modern GWASs generate numerous IVs suitable for MR analysis. For instance, the latest human height GWAS identified over 11,000 independent association signals [4], enhancing the power and accuracy of MR estimation.

However, many current MR analyses are challenged by a potentially severe confounding factor known as horizontal pleiotropy [5]. Horizontal pleiotropy violates the exclusive restriction assumption underlying the MR principle, casting doubt on the validity of MR analysis [6-8]. This issue becomes more pronounced as the number of IVs increases, as chance can lead to an increase in shared heritability factors between exposure and outcome. Therefore, effective correction for confounding due to horizontal pleiotropy is essential, especially in large sample settings.

Considerable efforts in the literature have addressed the pleiotropic effect and sought to mitigate its adverse impact on MR analysis [9, 10]. In a two-sample setting, multiple popular statistical methods accommodating pleiotropic IVs have been proposed and widely utilized [11-37]. However, under certain conditions, some methods may fail to adequately correct for horizontal pleiotropy or accurately detect true causal effects [9].

In this study, with the aim of conducting robust and accurate MR studies in the presence of horizontal pleiotropy, we introduce a novel statistical method. This method explicitly distinguishes causal effect from pleiotropic effect and estimates both within the maximum likelihood framework. Through simulation studies, we demonstrate that our proposed method effectively corrects for pleiotropic effects across various confounding scenarios while maintaining statistical power at a comparable level. As an application, we investigate the bidirectional causal effects of four lipid traits and appendicular lean mass (ALM) using data from the UK biobank (UKB) cohort and summary statistics from the Global Lipids Genetics Consortium (GLGC) [38]. Finally, we conduct a randomized controlled experiment in mice to address controversial findings revealed by our proposed method compared to alternative methods.

## Results

### Outline of the proposed method

The diagram illustrating the proposed method is presented in **Figure 1**. In this context, we consider the causal effect, denoted as *r*, of a continuous exposure trait *X* on a continuous outcome trait *Y*. Our objective is to estimate this causal effect using a collection of independent IVs that exhibit an association with *X*. These IVs are categorized into three distinct types based on their pleiotropic effects on the outcome and the correlation of these pleiotropic effects with the IV-exposure effects at corresponding IVs:

**Figure 1.**
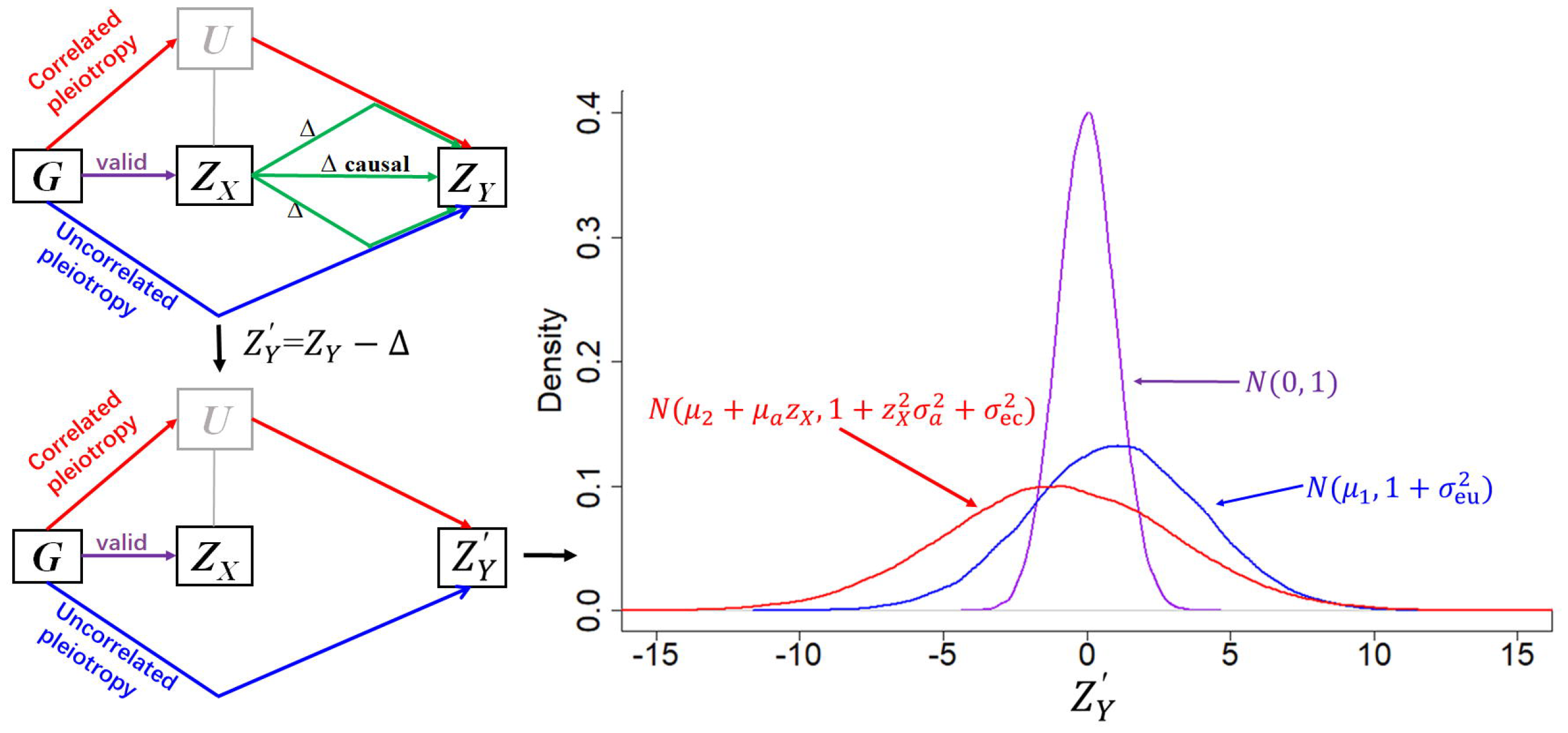
Diagram of the proposed method. Four elements are involved: IV *G*, exposure *X*, outcome *Y*, and confounder *U. X* and *Y* are represented by their association z-scores with *G*. There are three potential pathways from *G* to *Y*, depending on *G*’s pleiotropic status: valid (purple), uncorrelated pleiotropic (blue) and correlated pleiotropic (red). The causal effect Δ (green) exists in all pathways. After removing the causal effect Δ from *z*_*Y*_ (lower panel), the residual outcome z-score 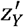 contains information on pleiotropic effects only. Depending on in which pathway *G* is, 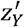 has a pathway-specific distribution (right panel), which is used to differentiate valid and pleiotropic IVs.

1. Valid IVs: These IVs do not exert horizontal pleiotropic effects on the outcome. They serve as reliable indicators of the exposure’s effect on the outcome without introducing additional confounding factors.
2. Uncorrelated pleiotropic IVs: This category comprises IVs that exhibit horizontal pleiotropic effects on the outcome. However, these effects are not correlated with the corresponding IV-exposure effects.
3. Correlated pleiotropic IVs: In this category, IVs demonstrate horizontal pleiotropic effects on the outcome that are correlated with the IV-exposure effects. This correlation violates the Instrument Strength Independent of Direct Effect (INSIDE) assumption [39].

In our analysis, we used genetic association z-scores, denoted as *z*_*X*_ and *z*_*Y*_, to estimate *r*. The outcome *z*_*Y*_ follows a normal distribution with a mean *μ*_γ_ and a variance of one, i.e. *Z*_*Y*∼_*N*(*μ*_γ,_1), where *μ*_γ_ represents the effect of the IV on the outcome. The causal effect of *X* on *Y* can be interpreted as shifting the mean *μ*_γ_ by an IV-specific offset denoted as 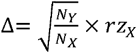, where *N* and *N* are the sample sizes of the exposure and outcome, respectively. Importantly, this shift applies uniformly across all IVs regardless of whether they are valid or pleiotropic. Consequently, if we subtract this offset Δ from *z*_*Y*_, we obtain a residual z-score 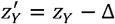 This residual z-score no longer contains any information about the causal effect.

We formulated 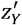 at different types of IVs to follow distinct distributions.

1. Valid IVs: 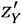 at valid IVs asymptotically follows a standard normal distribution, i.e.,

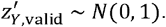

2. Uncorrelated pleiotropic IVs: At uncorrelated pleiotropic IVs, 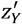 follows a normal distribution with a fixed mean parameter *μ*_1_ and an enlarged variance, i.e.,

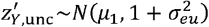

where *μ*_1_ and 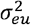 represent the mean and variance of the uncorrelated pleiotropic effects, respectively.

3. Correlated pleiotropic IVs: 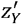 at correlated pleiotropic IVs follows another normal distribution. This distribution is characterized by a mean parameter related to *z*_*X*_ and an enlarged variance, i.e.,

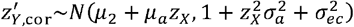

where *μ*_*a*_ and 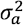 represent the mean and variance of the pleiotropic correlation, respectively. *μ*_2_ and 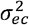 represent the mean and variance of the residual pleiotropic effects after adjusting for the pleiotropic correlation.

These formulations allow us to account for the different distributions of residual z-scores at various types of IVs, thereby capturing the distinct effects of horizontal pleiotropy on the outcome.

Given the presence of multiple independent IVs, we employed the maximum likelihood approach to estimate the parameters from the data. We assumed that the proportions of valid IVs, uncorrelated pleiotropic IVs, and correlated pleiotropic IVs are (1-*τ*), *τ* (1*-ρ*), and *τ ρ*, respectively. Here, *τ* and *ρ*, both fallowing within the range [0,1], represent the proportion of total pleiotropic IVs and the relative proportion of correlated pleiotropic IVs, respectively. Our model contains nine parameters in total: 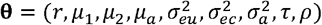. We constructed a likelihood function of **θ** as follows

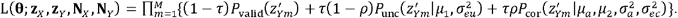

Maximizing L with respect to **θ** provides the maximum likelihood estimate of **θ**. From this estimate, we can derive both the estimated causal effect, denoted as 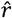, and the distribution of pleiotropic effects.

To assess the statistical significance of the estimates, we employed the likelihood ratio test (LRT) approach. Specifically, we examined the significance of 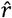 using a one-degree-of-freedom (df) chi-squared test. This test compares the maximized likelihood value with that obtained for a reduced likelihood function under the setting *r*=0. Additionally, we evaluated the existence of pleiotropic IVs using an 8-df chi-squared test. This test compares the maximized likelihood value with that obtained for another reduced likelihood function under the setting *τ*=0, in which *r* is the only model parameter.

### The distribution of 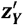

The distribution of 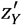 is central to our proposed method, <u>M</u>endelian <u>R</u>andomization analysis using <u>Z</u>-scores (MRZ), as it forms the basis for distinguishing between valid and pleiotropic IVs. Among the two parameters shaping the distribution of 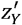, variance plays a crucial role in distinguishing between these categories. Through simulations, we investigated the distribution of 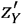 and evaluated the impact of three key factors on its variance: the sample size of the outcome (*N*_*Y*_), the variance 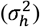 of the pleiotropic effect 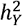, and the frequency of positive pleiotropic IVs (*f*_*p*_). Here, 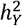 was defined as the IV-attributable portion of outcome variance, that is, IV-specific outcome heritability. As expected, 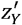 at valid IVs consistently conforms to a standard normal distribution (**Figure 2**). Conversely, at pleiotropic IVs, the variance of 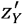 exceeds one and escalates with increasing *N*_*Y*_ and/or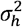. Specifically, when all pleiotropic IVs align in the same direction as the IV-exposure effects (*f*_*p*_=1), the variance of 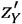 aligns well with theoretical expectations, which is 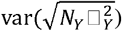. This variance amplifies when pleiotropic IVs consist of a mixture of positive and negative effects, reaching its maximum when the positive and negative pleiotropic IVs are evenly balanced (*f*_*p*_=0.5).

**Figure 2.**
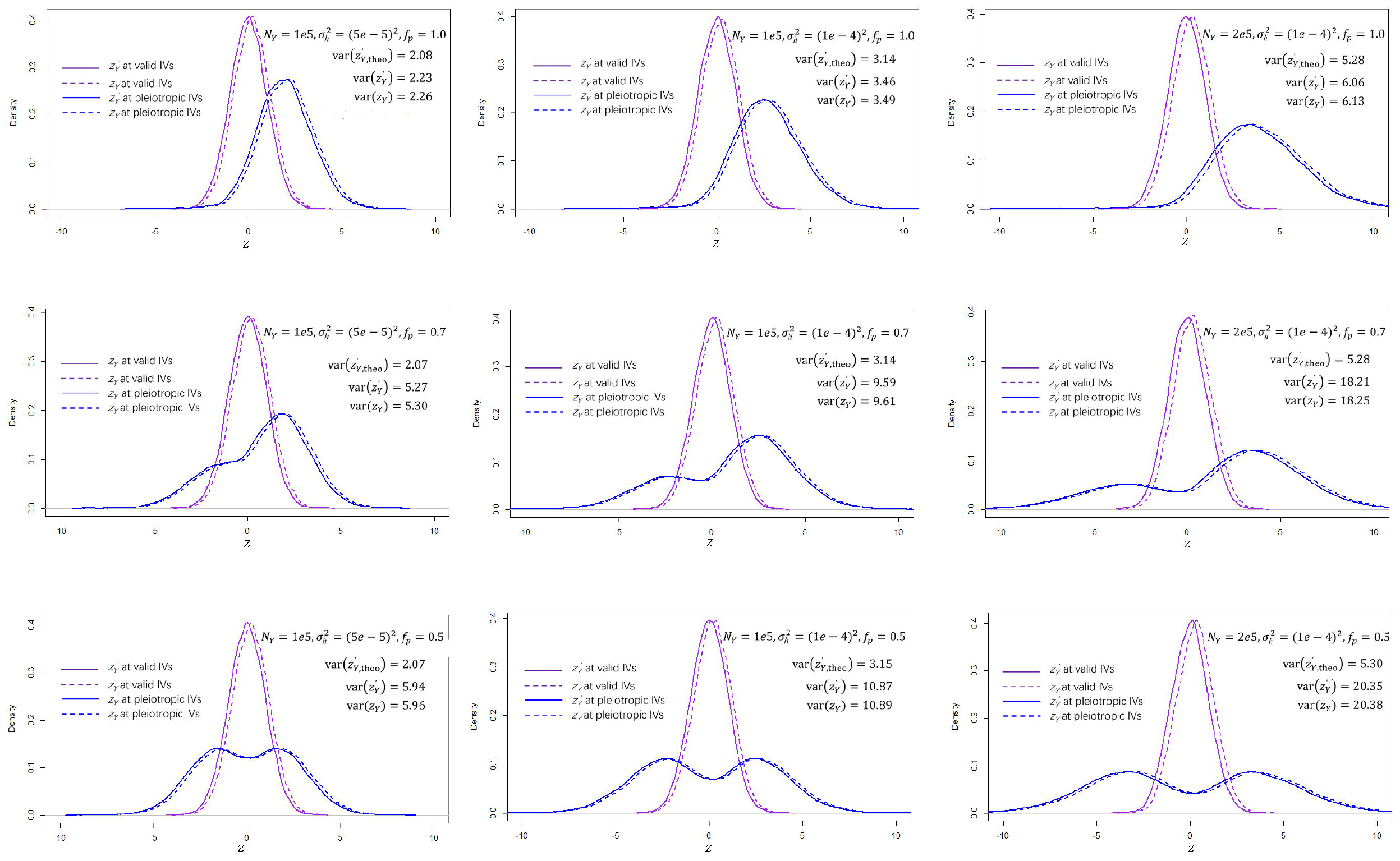
The distribution of outcome z-scores at valid and pleiotropic IVs. The influence of three factors on the distribution of outcome z-score was evaluated: outcome sample size (*N*_*Y*_*=*1e5 or 2e5), the variance of IV-specific heritability to the outcome 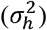, and the proportion of positive pleiotropic IVs (*fp*=0.5, 0.7 or 1.0). The IV-specific heritability 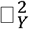 was drawn from an exponential distribution with a mean 5e-5 or 1e-4 so that its variance 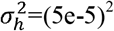 or (1e-4)^2^. The exposure sample size was set to be *N*_*X*_ =2e5. The causal effect of *X* on *Y* was set to be *r*=0.05. *z*_*Y*_ is IV-outcome z-score. 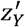 is the residual of *z*_*Y*_ after removing the causal effect. 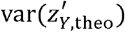 is the theoretical variance assuming *fp*=1.0. At valid IVs, both *z*_*Y*_ and 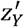 conform to a standard normal distribution. At pleiotropic IVs, the variance of 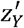 exceeds one and escalates with increasing *N*_*Y*_ and/or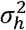. When *f*_*p*_=1.0, the variance of 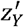 aligns well with 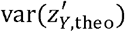, and amplifies when *f*_*p*_<1.0, peaking when the positive and negative pleiotropic IVs are evenly balanced (*f*_*p*_=0.5). *z*_*Y*_ has a similar distribution to 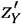.

Our simulations also revealed a striking resemblance between the distributions of 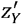 and *z*_*Y*_. This similarity arises from the positive nature of all *z*_*X*_ by definition, leading to a consistent shift of *z*_*Y*_ towards 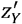 in the same direction across all IVs. Another fact reinforcing this similarity is the typically modest magnitude of the shift Δ, attributed to the small values of *r* in most real applications (e.g., <0.2). Consequently, we approximated the variance of 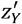 by studying *z*_*Y*_ as if no causal effect were present. By assuming an exponential distribution for pleiotropic effects, we estimated the distribution’s sole parameter [40]. Subsequently, we estimated the variance of 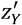 through Monto-Carlo sampling of 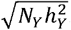. While this estimate assumes all pleiotropic effects align in the same direction and may thus underestimate the true variance, it provides a practical lower boundary from which the optimization of the likelihood function starts to work.

### Detecting pleiotropic effects

When all IVs are valid, MRZ does not detect any pleiotropy in both null and causal simulations (*P*<0.05). Notably, it is more conservative than MRPRESSO [19] and MREGGER [22], both of which identify pleiotropy and horizontal pleiotropy in approximately 5% of iterations.

In scenarios where pleiotropic IVs are present, both MRZ and MRPRESSO demonstrate remarkable efficacy, presenting 100% power in detecting pleiotropy even when the proportion of pleiotropic IVs is as low as 10% (**Figure 3A**). Conversely, MREGGER exhibits significantly lower power in discerning the directionality of pleiotropic effects. Even at the highest proportion (60%), MR-EGGER’s power rate is only 55.5%.

**Figure 3.**
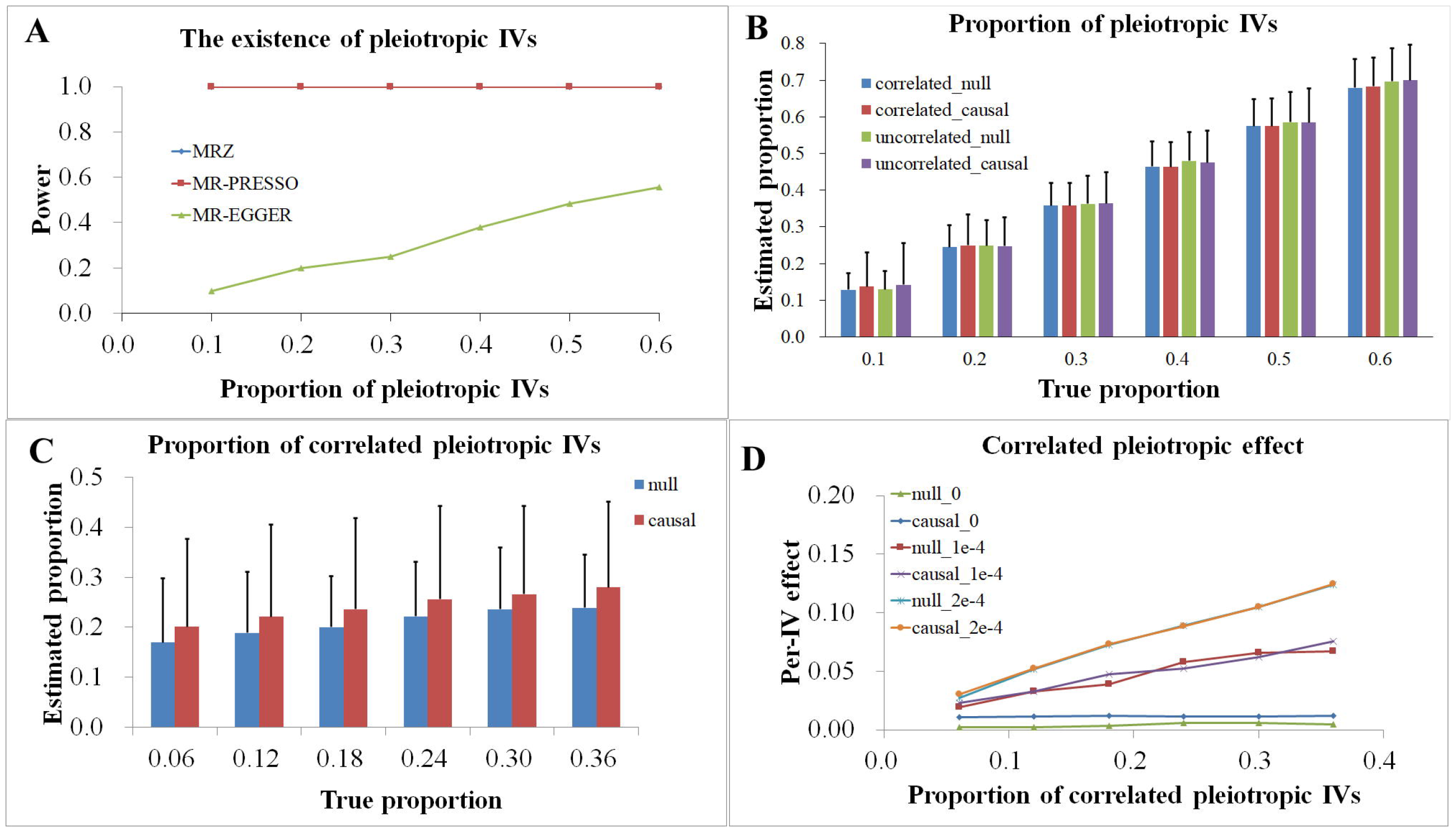
The performance of MRZ in detecting pleiotropic effects. A total of 200 IVs and 1000 iterations were simulated. **A**, The proportion of pleiotropic IVs ranges from 10%-60%. The relative proportion of correlated pleiotropic IVs is 50%. The power of detecting pleiotropy was declared at *α* =.0.05 level. **B**, Presented is the mean estimated proportion of pleiotropic IVs. Error bar is its standard deviation. Four scenarios were simulated, correlated pleiotropic effects and no causal effect (correlated_null), correlated pleiotropic effects and causal effect (correlated_causal), uncorrelated pleiotropic effects and no causal effect (uncorrelated_null), and uncorrelated pleiotropic effects and causal effect (uncorrelated_causal). **C**, In this simulation, the proportion of pleiotropic IVs was fixed at 60%, while the relative proportion of correlated pleiotropic IVs ranges from 10%-60%. Both causal and null scenarios were simulated. Presented is the mean estimated proportion of correlated pleiotropic IVs. The error bar is its standard deviation. **D**, the per-IV correlated pleiotropic effect was defined as the product of the proportion of correlated pleiotropic IVs and the mean correlated pleiotropic effect. The IV-specific heritability to the confounder was drawn from an exponential distribution with a mean 1e-4, 5e-5, or 0, respectively. These different settings correspond to different magnitudes of correlated pleiotropic effects.

The proportion of pleiotropic IVs estimated by MRZ exhibits a high correlation with the actual proportion, irrespective of the presence of a causal effect or the correlation status of pleiotropic effects (**Figure 3B**). However, a slight overestimate of the mean proportion by a relative proportion of approximately 10% is observed across all scenarios.

In instances where correlated pleiotropic IVs are present, MRZ’s ability to accurately estimate their proportion is unsatisfactory (**Figure 3C**). This suggests that uncorrelated and correlated pleiotropic IVs are indistinguishable by MRZ. Nevertheless, the per-IV mean correlated pleiotropic effect, defined as *τρμ*_*a*_, exhibits a perfect linear relationship with the proportion of correlated pleiotropic IVs (**Figure 3D**). The slope of this linear trend remains consistent between null and causal simulations, indicating minimal influence of the causal effect on the trend. Furthermore, the slope increases with stronger correlated pleiotropic effects and diminishes when no correlated pleiotropic IVs are included. Therefore, MRZ demonstrates the capability to capture correlated pleiotropic effects by jointly modeling the proportion of correlated pleiotropic IVs and the magnitudes of their effects.

### Causal effect: type-I error rate

Through a series of null simulations, we evaluated the type-I error rate of MRZ in testing causal effect. For comparative purposes, we incorporated 14 existing two-sample MR methods into our analysis. These include MREGGER [22], IVW [23], weighted-median (W-median) [21], weighted-mode (W-mode) [20], MRPRESSO [19], contamination mixture (CMix) [16], MRMix [18], MRAID [11], MRcML [12], GRAPPLE [13], MRLASSO [17], MRRAPS [14], MRROBUST [17], and CAUSE [15]. It’s worth noting that there are other promising methods not included in our analysis, some of which are extensively discussed elsewhere [9].

In the absence of pleiotropic effects, MRZ, along with other methods, effectively maintains type-I error rates close to the desired level of 5% (**Table 1**). However, W-mode (0.2%) and CAUSE (0.1%) demonstrate conservativeness, exhibiting lower type-I error rates.

**Table 1.** Performance of various methods with all valid IVs.

| Method | $r=0.00$ | $r=0.05$ | | |
| --- | --- | --- | --- | --- |
|  | Type-I error | Power | Effect | ME |
| MRZ | 0.052 | 0.999 | 0.049 | 0.008 |
| CAUSE | 0.001 | 0.800 | 0.050 | 0.008 |
| W-mode | 0.002 | 0.104 | 0.050 | 0.033 |
| MREGGER | 0.049 | 0.950 | 0.049 | 0.011 |
| MRMix | 0.068 | 0.259 | 0.049 | 0.025 |
| IVW | 0.065 | 0.999 | 0.049 | 0.008 |
| W-median | 0.031 | 0.948 | 0.049 | 0.010 |
| GRAPPLE | 0.056 | 0.999 | 0.050 | 0.008 |
| MRcML | 0.067 | 0.999 | 0.050 | 0.008 |
| RAPS | 0.057 | 0.999 | 0.050 | 0.008 |
| MRROBUST | 0.061 | 0.999 | 0.049 | 0.008 |
| MRLASSO | 0.073 | 0.998 | 0.049 | 0.008 |
| CMix | 0.054 | 0.998 | 0.049 | 0.008 |
| MRPRESSO | 0.050 | 0.998 | 0.049 | 0.008 |
| MRAID | 0.047 | 0.997 | 0.049 | 0.008 |

Notes: A total of 1,000 iterations were simulated. In each iteration, the sample size for both exposure and outcome samples was set to be 200,000. A total of 200 IVs were simulated, all of which are valid having no pleiotropic effect on the outcome. The 200 IVs explain approximately a portion of 5.8% of the exposure variance. Causal effect size was set to be $r=0.00$ (null) or $r=0.05$ (causal). Effect, mean estimated effect size across the 1,000 iterations; ME, mean error, defined as the absolute error of the estimated effect size from the true effect size $r=0.05$ .

In scenarios where pleiotropic effects are present, MRZ consistently maintains the correct type-I error rate across various proportions of pleiotropic IVs, irrespective of whether the pleiotropic effects are uncorrelated or correlated (**Table 2**). This robust performance also holds true under balanced pleiotropic effects (**Supplemental Table 1**). Among alternative methods, W-mode and CAUSE tend to be overly conservative, resulting in significantly reduced type-I error rates in all scenarios (**Table 2**). MREGGER shows validity only when pleiotropic effects are uncorrelated but exhibits an inflated type-I error rate of up to 28.1% when the pleiotropic effects are correlated. MRMix generally performs well under low to modest proportions of pleiotropic IVs but shows an inflated error rate reaching up to 21.6% under higher proportions. All the other alternative methods have inflated type-I error rates that increase with increased proportion of pleiotropic IVs, whereas the inflation is more severe under correlated pleiotropy than under uncorrelated pleiotropy. The type-I error rates for certain methods can reach nearly 100% in some extreme scenarios, rendering them invalid at all under such conditions. Even when the pleiotropic effects are balanced, the inflation of type-I error rates is still observed for most alternative methods, especially in correlated pleiotropic scenarios (**Supplemental Table 1**).

**Table 2.** Type-I error rates of various methods under pleiotropic effects.

| PleiotropicIVs(%) | MRZ | CAUSE | W-mode | EGGER | MRMix | MRAID | IVW | W-median | CMix | GRAPPLE | MRcML | MRROBUST | MRLASSO | RAPS | MRPRESSO |
| --- | --- | --- | --- | --- | --- | --- | --- | --- | --- | --- | --- | --- | --- | --- | --- |
| Uncorrelated pleiotropy |  |  |  |  |  |  |  |  |  |  |  |  |  |  |  |
| 10 | 0.05 | 0.00 | 0.00 | 0.06 | 0.03 | 0.06 | 0.15 | 0.05 | 0.07 | 0.04 | 0.12 | 0.08 | 0.10 | 0.08 | 0.08 |
| 20 | 0.04 | 0.00 | 0.00 | 0.06 | 0.06 | 0.08 | 0.30 | 0.07 | 0.12 | 0.13 | 0.26 | 0.12 | 0.16 | 0.16 | 0.19 |
| 30 | 0.04 | 0.00 | 0.00 | 0.05 | 0.05 | 0.14 | 0.47 | 0.12 | 0.24 | 0.31 | 0.42 | 0.24 | 0.30 | 0.31 | 0.49 |
| 40 | 0.07 | 0.01 | 0.01 | 0.06 | 0.09 | 0.22 | 0.60 | 0.21 | 0.40 | 0.54 | 0.61 | 0.47 | 0.51 | 0.50 | 0.66 |
| 50 | 0.05 | 0.01 | 0.01 | 0.06 | 0.10 | 0.31 | 0.69 | 0.30 | 0.53 | 0.68 | 0.75 | 0.69 | 0.71 | 0.61 | 0.83 |
| 60 | 0.04 | 0.02 | 0.01 | 0.05 | 0.22 | 0.44 | 0.77 | 0.45 | 0.73 | 0.81 | 0.87 | 0.83 | 0.89 | 0.73 | 0.92 |
| Correlated pleiotropy |  |  |  |  |  |  |  |  |  |  |  |  |  |  |  |
| 10 | 0.03 | 0.00 | 0.00 | 0.10 | 0.02 | 0.04 | 0.32 | 0.03 | 0.05 | 0.03 | 0.10 | 0.05 | 0.07 | 0.06 | 0.11 |
| 20 | 0.06 | 0.00 | 0.00 | 0.12 | 0.04 | 0.08 | 0.62 | 0.08 | 0.12 | 0.25 | 0.21 | 0.11 | 0.19 | 0.20 | 0.25 |
| 30 | 0.05 | 0.01 | 0.00 | 0.16 | 0.05 | 0.10 | 0.81 | 0.17 | 0.20 | 0.62 | 0.39 | 0.24 | 0.38 | 0.51 | 0.69 |
| 40 | 0.04 | 0.01 | 0.01 | 0.20 | 0.05 | 0.15 | 0.93 | 0.30 | 0.34 | 0.87 | 0.57 | 0.59 | 0.67 | 0.82 | 0.91 |
| 50 | 0.05 | 0.02 | 0.01 | 0.24 | 0.08 | 0.25 | 0.98 | 0.49 | 0.54 | 0.97 | 0.74 | 0.93 | 0.89 | 0.94 | 0.95 |
| 60 | 0.05 | 0.02 | 0.03 | 0.28 | 0.18 | 0.40 | 0.99 | 0.67 | 0.70 | 0.99 | 0.88 | 0.98 | 0.97 | 0.98 | 1.00 |

Notes: A total of 200 IVs and 1,000 iterations were simulated. Sample size for both exposure and outcome samples was set to be 2e5. The 200 IVs explain approximately a portion of 5.8% of the exposure variance. In the case of uncorrelated pleiotropy, 70% of all pleiotropic IVs were randomly selected to have positive pleiotropic effects while the remaining 30% pleiotropic IVs were simulated to have negative pleiotropic effects. In the case of correlated pleiotropy, 50% of pleiotropic IVs were simulated to be correlated with the corresponding IV-exposure effects. The causal effect $r$ was set to be 0. The significance threshold was set to 0.05.

Further simulation studies involving a smaller set of 100 IVs (**Supplemental Table 2**) or a larger set of 500 IVs (**Supplemental Table 3**) reaffirm the validity of MRZ. Among alternative methods, W-mode and CAUSE exhibit inflated type-I error rates under high proportions of pleiotropic IVs when the number of IVs is 100 and 500, respectively, rendering them invalid (**Supplemental Tables 2-3**). Therefore, our simulations reveal MRZ’s efficacy in rectifying horizontal pleiotropy across diverse confounding scenarios, where existing methods often lose validity.

Notably, MRZ demonstrates strong robustness against sample overlap due to its effective control of horizontal pleiotropy. It maintains a valid type-I error rate even when exposure and outcome samples completely overlap (**Supplemental Table 4**), making it suitable for scenarios involving a single biobank-scale dataset such as the UKB cohort.

### Causal effect: power and effect size

We conducted a series of causal simulations to examine the power and effect size of various methods. In scenarios without pleiotropic effects, nearly all alternative liberal methods—those prone to type-I error under pleiotropic scenarios—demonstrate remarkably high power rates almost reaching 100% (**Table 1**). MRZ notably achieves a power rate of 99.9%, positioning it among the most powerful methods. Conversely, W-mode exhibits the lowest power rate (10.4%), followed by MRMix (25.9%) and CAUSE (80.0%). Regarding effect size estimation, most methods, including MRZ, estimate mean effect sizes close to the true value of 0.050. MRZ displays one of the lowest mean errors (MEs, 0.008) among all methods. In contrast, W-mode (0.033) and MRMix (0.025) exhibit larger variations.

In the presence of horizontal pleiotropy, MRZ exhibits a decline in power as the proportion of pleiotropic IVs increases, as expected. This decline is similar in both positive and negative causal effect settings (**Supplemental Table 5**), indicating minimal impact of pleiotropic effects on MRZ’s power across diverse scenarios. Even with the highest proportion of 60% pleiotropic IVs, MRZ’s power rate maintains between 40%-58%.

Alternative methods demonstrate scenario-dependent performance, lacking a universal trend across all settings. The two overly conservative methods CAUSE and W-mode, along with MREGGER, MRMix and MRAID, generally exhibit declining power with increased proportion of pleiotropic IVs (**Supplemental Table 5**). For other alternative methods, two distinct trends are observed depending on the relative directions of the causal effect and the pleiotropic effects. When they align, these methods maintain high power rates at nearly 100% at all proportions of pleiotropic IVs. Conversely, when they oppose, these methods experience rapid decline in power rates with increased proportion of pleiotropic IVs (**Supplemental Table 5**). These conflicting trends highlight potential confounding effects of uncorrected pleiotropy on power estimation.

To ensure a fair comparison of power rates among methods despite varying type-I error rates, we corrected each method’s raw power rate by its type-I error rate at the corresponding null setting, assuming a correct type-I error rate of 5%. Like MRZ, all alternative methods have a decrease in their corrected power rates when the proportion of pleiotropic IVs increases (**Figure 4**). Among the methods, MRZ generally maintains the highest corrected power rate across various proportions of pleiotropic IVs. This improvement is particularly notable when the causal effect opposes the pleiotropic effects, in which cases uncorrected pleiotropic effects counteract or even reverse the true causal effect, resulting in a severe loss of power for alternative methods.

**Figure 4.**
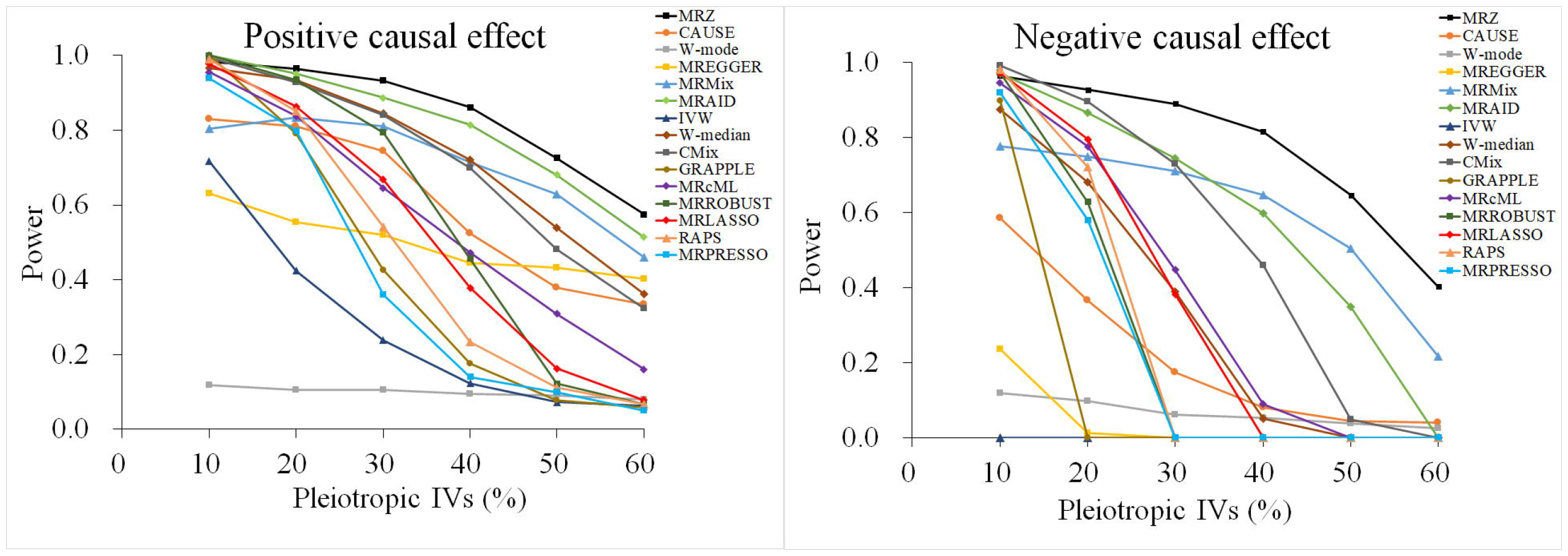
Corrected power rate of various methods for testing causal effect. A total of 200 IVs were simulated. The sample sizes for both exposure and outcome samples were 2e5. The 200 IVs explained approximately a portion of 5.8% of the exposure variance. A positively dominated pleiotropic setting was simulated, in which 70% pleiotropic IVs were simulated to have positive pleiotropic effects while the remaining 30% were simulated to have negative pleiotropic effects. In addition, half pleiotropic IVs were simulated to be correlated with the corresponding IV-exposure effects. The causal effect *r* was set to be 0.05 (positive effect) or -0.05 (negative effect). The statistical significance was declared at *α* =.0.05 level. Raw power rate was corrected by calibrating it with the type-I error rate estimated at the corresponding null condition, using the equation

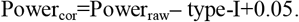

Uncorrected pleiotropic effects not only affect the power to detect the causal effect but also influence estimated effect size. MRZ’s estimated effect sizes align with the true value (0.05 or -0.05) across all scenarios (**Figure 5)**, regardless of the proportion of pleiotropic IVs or the direction of the causal effect. Conversely, for all alternative methods, including W-mode and CAUSE, distinct trends are observed based on the relative directions of the causal effect to pleiotropic effects. When the directions align, mean effect sizes of all alternative methods tend to increase with the proportion of pleiotropic IVs, whereas they decrease when the directions oppose. At the highest proportion of 60% pleiotropic IVs, the decrease is so severe that the estimated effect sizes from all alternative methods are opposite to the true size.

**Figure 5.**
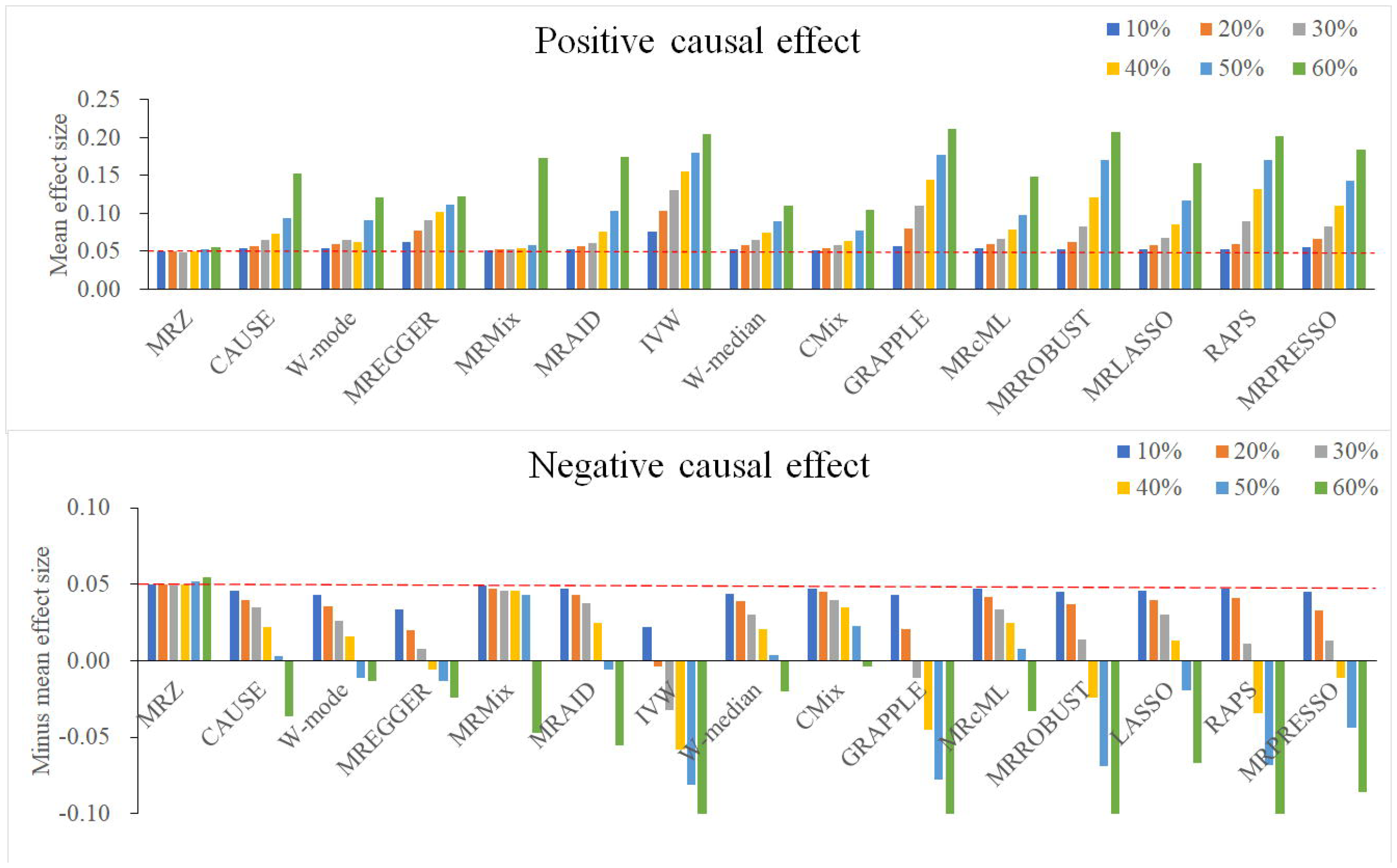
Estimated causal effect size of various methods. The causal effect *r* was set to be 0.05 (positive effect) or -0.05 (negative effect). The mean effect size across 1,000 iterations is presented.

Additional simulation studies with 100 and 500 IVs (**Supplemental Figure 1**) reveal an increased trend of MRZ’s power rate with an increasing number of IVs, underscoring the critical importance of including a substantial number of IVs for robust and powerful causal inference.

When modeling no pleiotropic effects, the proposed test statistic *T*_1_ closely approximates the causal effect size estimated by the IVW test across all simulated scenarios (**Supplemental Figure 2)**. This suggests that the IVW test provides a reasonable anchor for MRZ to adjust its estimated effect size when analyzing un-standardized summary statistics, such as case-control data.

### Running time

The computation process of MRZ primarily focuses on optimizing the likelihood function twice: once for the alternative hypothesis and once for the null hypothesis. Despite involving a high dimension of nine parameters, the derivative-free optimization algorithm *nmkb* that we employed offers an efficient solution. MRZ completes the optimization within seconds even on datasets containing 500 IVs. However, it’s important to acknowledge that there is no assurance that the optimization algorithm will converge to its global maximum under such a high-dimensional parameter space. Therefore, it’s recommended to conduct repeated optimizations with varying initial parameter settings to enhance the likelihood of obtaining a robust solution. In this study, a total of 20 repeats were performed to ensure the reliability of the results.

### Real data analysis

As an application, we conducted a bidirectional MR study examining the relationship between ALM and four circulating lipid traits (high-density lipoprotein cholesterol (HDL-C), low-density lipoprotein cholesterol (LDL-C), total cholesterol (TC), and triglycerides (TG)). We utilized two data sources: the UKB internal cohort and the summary statistics released by GLGC.

In the UKB-internal analysis, we randomly divided the entire UKB cohort into two independent sub-samples (UKB_S1 and UKB_S2). For each exposure-outcome pair (e.g., ALM-TC), one sub-sample served as the exposure sample while the other served as the outcome sample. This process was repeated twice by reversing the roles of the two sub-samples so that two independent sets of summary statistics were generated [41]. In the UKB-GLGC joint analysis, we used the entire UKB cohort to generate GWAS summary statistics for ALM, while GLGC data (excluding UKB participants) provided summary statistics for the lipid traits. Consequently, we generated three datasets that are mutually independent in exposure and/or outcome, allowing for cross-validation of results.

To assess robustness against sample overlap, we conducted an MR analysis using both exposure and outcome summary statistics derived from the entire UKB cohort, resulting in complete sample overlap.

Observational analyses reveal significant correlations between raw ALM values and all lipid traits in the UKB cohort, and these correlations persist after adjusting for age and sex in both ALM and lipid traits (**Supplemental Table 6**). In the MR settings, the number of eligible IVs ranges from 85 to 719 across all exposure-outcome pairs (**Supplemental Tables 7-8**). Strong IV-exposure associations are observed, with *R*^2^ values ranging from 0.05 to 0.10 and *F*-statistics ranging from 52.1 to 188.8.

At a significance level of 6.25×10^−3^ (0.05/(2×4)), forward MRZ analysis identified a negative association between ALM and both TC and LDL-C (**Table 3**). This association is statistically significant in all three datasets for both traits, strengthening the evidence of causality. The estimated effect sizes are consistent across datasets for both TC (−0.079, -0.083 and -0.074) and LDL-C (−0.054, -0.063 and -0.037). Notably, the proportion of pleiotropic IVs was estimated to be modest to high for both traits, being around 30% and 40% in the UKB-internal datasets (261-268 IVs).

**Table 3.**
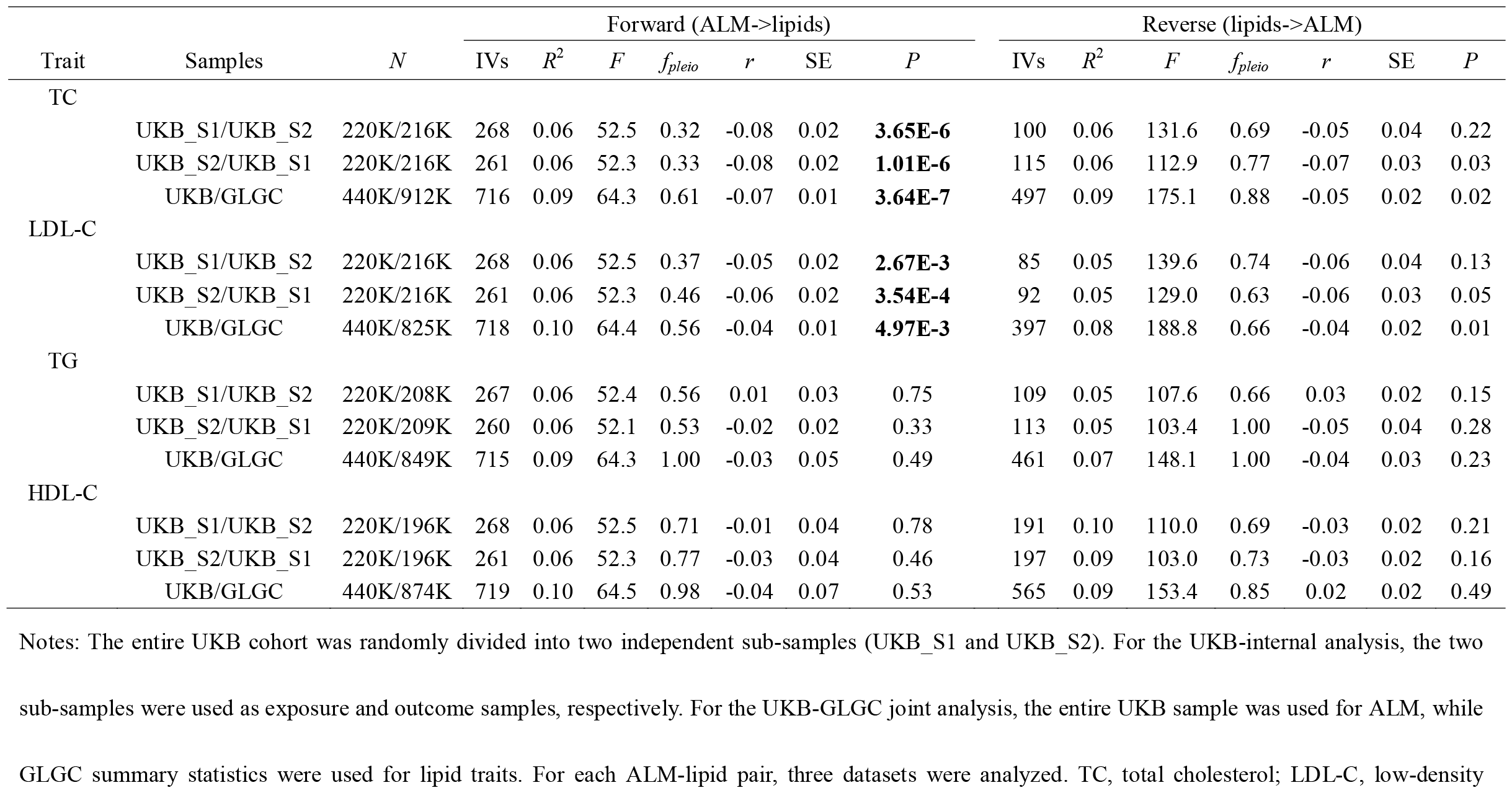
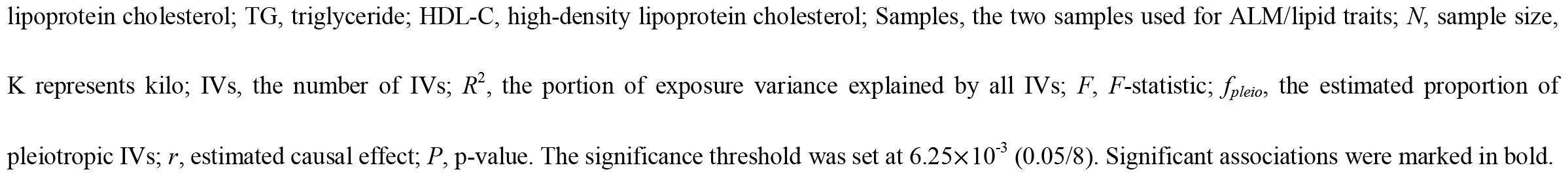
Bidirectional causal effects of ALM and lipid traits identified by MRZ.

| Trait | Samples | *N* | Forward (ALM->lipids) |  |  |  |  |  |  | Reverse (lipids->ALM) |  |  |  |  |  |  |
| --- | --- | --- | --- | --- | --- | --- | --- | --- | --- | --- | --- | --- | --- | --- | --- | --- |
|  |  |  | IVs | *R*<sup>2</sup> | *F* | *f*<sub>pleio</sub> | *r* | SE | *P* | IVs | *R*<sup>2</sup> | *F* | *f*<sub>pleio</sub> | *r* | SE | *P* |
| TC | UKB_S1/UKB_S2 | 220K/216K | 268 | 0.06 | 52.5 | 0.32 | -0.08 | 0.02 | **3.65E-6** | 100 | 0.06 | 131.6 | 0.69 | -0.05 | 0.04 | 0.22 |
|  | UKB_S2/UKB_S1 | 220K/216K | 261 | 0.06 | 52.3 | 0.33 | -0.08 | 0.02 | **1.01E-6** | 115 | 0.06 | 112.9 | 0.77 | -0.07 | 0.03 | 0.03 |
|  | UKB/GLGC | 440K/912K | 716 | 0.09 | 64.3 | 0.61 | -0.07 | 0.01 | **3.64E-7** | 497 | 0.09 | 175.1 | 0.88 | -0.05 | 0.02 | 0.02 |
| LDL-C | UKB_S1/UKB_S2 | 220K/216K | 268 | 0.06 | 52.5 | 0.37 | -0.05 | 0.02 | **2.67E-3** | 85 | 0.05 | 139.6 | 0.74 | -0.06 | 0.04 | 0.13 |
|  | UKB_S2/UKB_S1 | 220K/216K | 261 | 0.06 | 52.3 | 0.46 | -0.06 | 0.02 | **3.54E-4** | 92 | 0.05 | 129.0 | 0.63 | -0.06 | 0.03 | 0.05 |
|  | UKB/GLGC | 440K/825K | 718 | 0.10 | 64.4 | 0.56 | -0.04 | 0.01 | **4.97E-3** | 397 | 0.08 | 188.8 | 0.66 | -0.04 | 0.02 | 0.01 |
| TG | UKB_S1/UKB_S2 | 220K/208K | 267 | 0.06 | 52.4 | 0.56 | 0.01 | 0.03 | 0.75 | 109 | 0.05 | 107.6 | 0.66 | 0.03 | 0.02 | 0.15 |
|  | UKB_S2/UKB_S1 | 220K/209K | 260 | 0.06 | 52.1 | 0.53 | -0.02 | 0.02 | 0.33 | 113 | 0.05 | 103.4 | 1.00 | -0.05 | 0.04 | 0.28 |
|  | UKB/GLGC | 440K/849K | 715 | 0.09 | 64.3 | 1.00 | -0.03 | 0.05 | 0.49 | 461 | 0.07 | 148.1 | 1.00 | -0.04 | 0.03 | 0.23 |
| HDL-C | UKB_S1/UKB_S2 | 220K/196K | 268 | 0.06 | 52.5 | 0.71 | -0.01 | 0.04 | 0.78 | 191 | 0.10 | 110.0 | 0.69 | -0.03 | 0.02 | 0.21 |
|  | UKB_S2/UKB_S1 | 220K/196K | 261 | 0.06 | 52.3 | 0.77 | -0.03 | 0.04 | 0.46 | 197 | 0.09 | 103.0 | 0.73 | -0.03 | 0.02 | 0.16 |
|  | UKB/GLGC | 440K/874K | 719 | 0.10 | 64.5 | 0.98 | -0.04 | 0.07 | 0.53 | 565 | 0.09 | 153.4 | 0.85 | 0.02 | 0.02 | 0.49 |

Notes: The entire UKB cohort was randomly divided into two independent sub-samples (UKB\_S1 and UKB\_S2). For the UKB-internal analysis, the two
sub-samples were used as exposure and outcome samples, respectively. For the UKB-GLGC joint analysis, the entire UKB sample was used for ALM, while
GLGC summary statistics were used for lipid traits. For each ALM-lipid pair, three datasets were analyzed. TC, total cholesterol; LDL-C, low-density
lipoprotein cholesterol; TG, triglyceride; HDL-C, high-density lipoprotein cholesterol; Samples, the two samples used for ALM/lipid traits; $N$ , sample size, $K$ represents kilo; IVs, the number of IVs; $R^2$ , the portion of exposure variance explained by all IVs; $F$ , $F$ -statistic; $f_{pleio}$ , the estimated proportion of pleiotropic IVs; $r$ , estimated causal effect; $P$ , p-value. The significance threshold was set at $6.25 \times 10^{-3}$ (0.05/8). Significant associations were marked in bold.

Forward MR analyses using MRZ did not detect significant associations between ALM and either HDL-C or TG in any of the datasets, suggesting no causal effects on both traits. Further reverse MR analyses did not identify significant associations for any lipid traits with ALM, indicating no reverse causal effect. Analysis on the completely overlapped whole UKB cohort yielded similar results (**Supplemental Table 9**), confirming MRZ’s robustness against sample overlap.

We compared the MRZ results with those from alternative methods. In the forward MR analyses, all alternative methods identified the same negative causal effect of ALM on TC in one or more datasets (**Supplemental Figure 3**). However, in the reverse MR analyses, all alternative methods revealed a same negative causal effect of TC on ALM in at least one dataset (**Supplemental Figure 3**). The results of mutual causality at the same direction are observed from alternative methods on the other three lipid traits too (**Supplemental Figures 4-6**). Observing mutually reinforcing causal effects at the same direction suggests a high likelihood of pleiotropy, making it difficult to definitively determine the true causal relationship. Therefore, leaving aside the true causal relationships, none of the alternative methods can produce results that are free of pleiotropic effects and that are self-validated across all datasets.

### Experimental validation

Given the unidirectional causal link identified by MRZ versus the disputed bidirectional link suggested by alternative methods, there are conflicting views on the causal effect of lipid traits on ALM. To address this controversy empirically, we conducted an *in vivo* randomized controlled experiment using a mouse model. The experiments involved intervention with TC and compared to normal controls. The results reveal a significant increase in circulating TC levels in mice fed TC for a period of 8 weeks (*N*=12) compared to normal controls (*N*=12, Wilcoxon rank test *P*=3.59 ×10^−5^, **Figure 6**), indicating successful implementation of the TC intervention. Additionally, significant changes are observed in HDL-C (*P*=1.24 ×10^−14^), LDL-C (*P*=5.98 ×10^−9^), and TG (*P*=5.98 ×10^−5^) levels.

**Figure 6.**
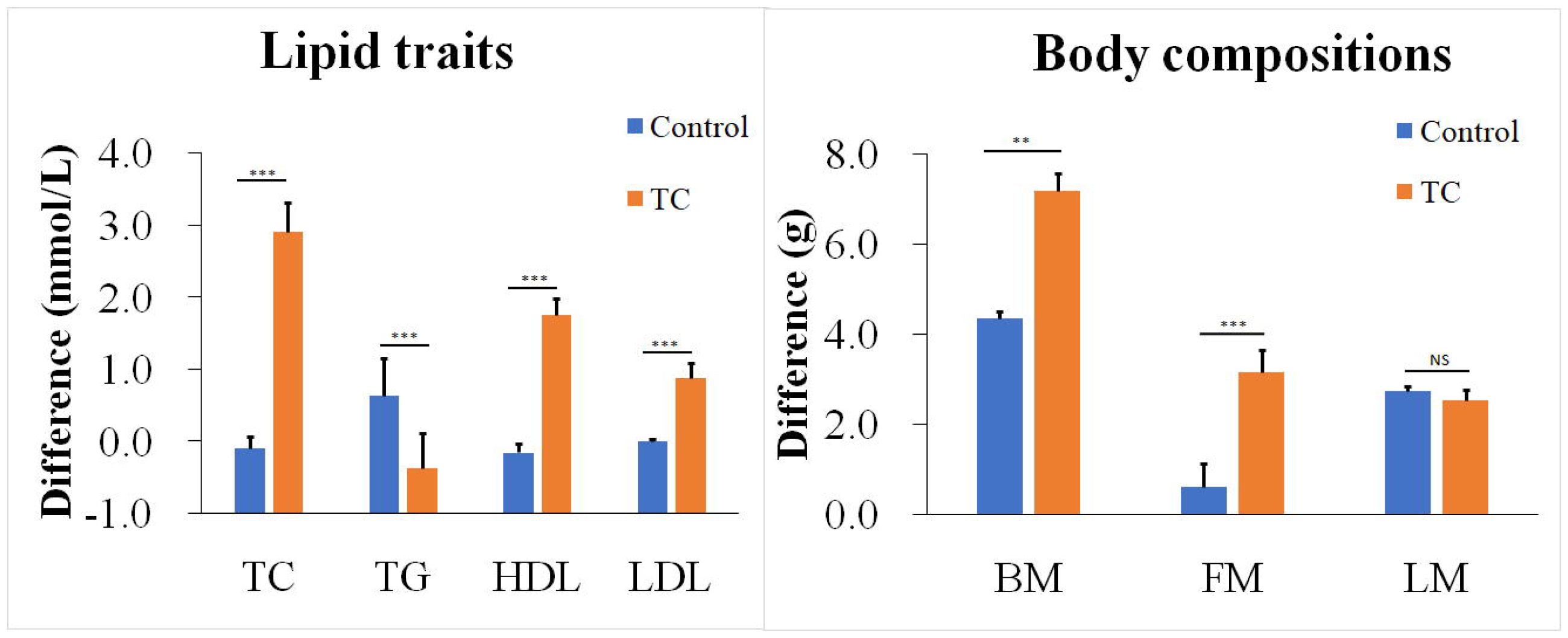
The randomized controlled experiment intervened by TC in mice. Male C57BL/6 mice were randomly allocated to control group (*N*=12) and intervention group (*N*=12), followed by intervention by TC diet in the TC group for 8 weeks. For each trait, the difference of measurement between the endpoint and the baseline was analyzed. The comparison was made between the control group and the intervention group using the Wilcoxon rank test. ***, *P*<0.001; **, *P*<0.001; NS, non-significant (*P*>0.05).

As the outcome, we observed a significant increase in total body mass (*P*=1.64 ×10^−3^) and fat body mass (*P*=1.03 ×10^−4^) in the TC intervention group. However, there is no significant difference in lean body mass (*P*=0.80), suggesting no causal effect of lipid traits on ALM. These findings are consistent with the results obtained from MRZ analyses, but differ from the conclusions drawn by most alternative methods. This highlights MRZ’s resilience against horizontal pleiotropic effects, to which existing methods are more susceptible.

## Discussion

Under the prevailing polygenic genetic architecture of complex traits, pleiotropy is anticipated to be a common occurrence [42]. While theoretical assertions suggest that perfectly balanced pleiotropic effects could cancel out bias [22], achieving such equilibrium is impractical. Current MR practices often employ sensitivity analyses like the MREGGER intercept test to evaluate pleiotropy balance [22]. However, our simulation studies revealed that the MREGGER intercept test lacks sufficient power to detect even substantial pleiotropic imbalances. Thus, relying solely on these methods cannot ensure that empirical MR analyses are conducted under balanced pleiotropic conditions. Even under balanced pleiotropy, our simulation studies demonstrated that the type-I error rate of certain methods may inflate as the proportion of pleiotropic IVs increases. The presence of correlated pleiotropic effects exacerbates this issue. Therefore, robust methods to address horizontal pleiotropy are crucial to ensure the validity of MR analyses.

Our proposed method, MRZ, categorizes IVs into three distinct groups reflecting potential pathways from IVs to the outcome, as depicted in the classical MR rationale diagram. By assuming a general normality distribution for underlying pleiotropic effects, we derived a precise distribution for IV-outcome z-scores specific to each category. In simulated scenarios encompassing diverse pleiotropic settings, MRZ demonstrates enhanced control of type-I error rate as well as more precise and powerful causal effect estimation than existing methods. Notably, the normality assumption assumed by MRZ was not met in our simulations. However, we did not observe inflated type-I error rates or imprecise effect estimates, demonstrating the robustness of MRZ against data distribution.

Multiple sophisticated models have been proposed to address horizontal pleiotropy in MR analyses, with some examples provided in this study. While these methods demonstrate excellent performance under specific conditions, our simulation studies reveal limitations when the number of IVs reaches the hundreds. In such scenarios, certain alternative methods struggle to effectively correct for pleiotropic effects. This challenge arises because pleiotropic effects are often systematic, meaning they are not simply averaged out with an increasing number of IVs. In fact, the presence of systematic pleiotropy can worsen as the number of IVs grows.

Among alternative methods, MRMix shares a similar strategy of categorizing IVs based on their potential causal effects [18]. However, MRMix utilizes four categories reflecting all combinations of the presence or absence of direct effects between IVs and both the exposure and the outcome. This classification allows MRMix to include IVs with no association to the exposure. In contrast, MRZ assumes all IVs to be associated with the exposure, aligning with the standard practice of screening and filtering IVs in empirical MR analyses to ensure this condition.

Another key distinction between MRZ and MRMix lies in the data modeling strategy. MRMix assumes both IV-exposure and IV-outcome effects are random variables following a bivariate normal distribution. Conversely, MRZ treats the IV-exposure effect as a fixed value and models only the IV-outcome effect as a random variable with a distribution conditional on the former. This approach in MRZ eliminates the requirement for a normality assumption on the IV-exposure effect, making it more robust to non-normality, a frequent characteristic in non-infinitesimal genetic models.

Another comparable method, CMix, employs distinct probability functions to model valid and pleiotropic IVs as well. Key disparities between MRZ and CMix include the following: 1) MRZ categorizes pleiotropic IVs into uncorrelated and correlated groups, offering a more detailed classification compared to CMix, which treats all pleiotropic IVs collectively; 2) Both methods assume a normal distribution for effect sizes of pleiotropic IVs. However, MRZ allows the mean parameter of this distribution to vary freely, while CMix constrains it to zero but allows for an expanded variance; 3) MRZ integrates uncertainty regarding IV validity into its likelihood function without explicitly designating each IV as valid or pleiotropic. Conversely, CMix explicitly assigns a label to each IV based on its estimated probability of being valid or pleiotropic under each parameter setting. We argue that incorporating IV uncertainty may provide valuable insights into the distribution of pleiotropic IVs. Our simulation studies demonstrate that MRZ can effectively estimate the proportion of pleiotropic IVs, thereby supporting its efficacy.

Skeletal muscle and circulating lipids play a critical role in regulating energy balance [43-45]. They are intricately linked and share common genetic background [46, 47]. Our real data analysis revealed a high degree of pleiotropy between ALM and all the lipid traits we examined. This pleiotropy poses a significant challenge for most existing MR methods. These methods tend to have inflated type-I error rates when the proportion of pleiotropic IVs is modest to high. As evidence, all alternative methods discovered a causal effect of TC on ALM in at least one dataset. However, this finding contradicted the results of controlled experiments where TC levels were directly manipulated. In contrast, MRZ yielded a different result that aligned with the experiment results. This highlights the importance of robust methods for accurate causal inference in MR studies, especially when pleiotropy is a major concern.

The observed causal effect of ALM on TC and LDL-C in our real data analysis is supported by nearly all alternative methods. Several plausible biological mechanisms could underlie this causality: 1) Metabolic rate and energy balance: Lean mass contributes significantly to basal metabolic rate [48, 49], which in turn can influence lipid metabolism and cholesterol levels. 2) Insulin sensitivity: Muscle tissue plays a crucial role in glucose metabolism and insulin sensitivity [50]. Higher lean mass is often associated with improved insulin sensitivity and glucose uptake [50], which in turn lead to changes in TC and LDL-C levels [51]; 3) Anti-inflammatory effects of muscle: Muscle secretes beneficial hormones such as interleukin-6 (IL-6) and irisin [52, 53], which have anti-inflammatory properties. These hormones can influence lipid metabolism and cholesterol levels [54, 55].

It should be notable that ALM is usually altered by other modifiable factors such as exercise and diet. In this case, they are in the same causal pathway, and it is unclear whether the changes in circulating TC and LDL-C are directly caused by changes in ALM or if ALM acts as a mediator of some modifiable factor. Further investigation is warranted to elucidate the biological mechanism underlying this causal relationship.

Certain limitations exist in the proposed method. Firstly, it does not account for certain biases common in MR analyses, such as weak instrument bias [56] and winner’s curse bias [57]. These biases can potentially be mitigated by implementing stringent IV filtering, such as applying a more rigorous significance threshold [57]. Secondly, optimizing a high-dimensional likelihood function poses challenges in converging to its global solution. To enhance the likelihood of reaching the global maximum, it is advisable to perform multiple optimizations with varying initial parameters. However, this approach increases computational burden and does not guarantee attainment of the global maximum. Thirdly, the proposed method requires both exposure and outcome sample sizes as input, unlike other methods that do not have this requirement. This necessity arises because z-scores are meaningful only within the context of a specific sample size. If exposure and outcome sample sizes are uniform so that their ratios are constant across all IVs, then the detailed sample sizes are indeed unnecessary because the estimated effect size remains unchanged after calibrating to the effect size estimated by the IVW test. However, if sample sizes vary across IVs, then providing this information is essential for unbiased estimation by the proposed method. Furthermore, in scenarios involving case-control studies with imbalanced case-to-control ratios, employing an effective sample size rather than a raw sample size is preferable for accurate estimation [58].

In summary, we have proposed a novel two-sample MR method that demonstrates robustness against horizontal pleiotropic effects, while also offering accuracy and power across a wide range of scenarios. By applying this method to investigate the relationship between ALM and lipid traits, we identified a negative causal effect of ALM on TC and LDL-C. Our proposed method serves as a valuable alternative to existing MR methods, particularly in the analysis of large-scale biobank datasets with numerous IVs. With its ability to provide reliable and precise causal inference, our method contributes to advancing the field of MR analysis and enhances our understanding of complex biological relationships.

## Online methods

### Basic model

We assumed a continuous exposure trait *X*, a continuous outcome trait *Y*, and a bi-allelic SNP *G* taking values between 0, 1, and 2. For ease of presentation, we assumed *X* and *Y* are standardized so that their variances are one. Assumed that *G* is associated with *X* and serves as its IV and that the associations of *G* with both *X* and *Y* were tested under an additive mode of inheritance.

The basic phenotype model for MR analysis was formulated as following

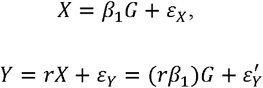

where *β*_1_ measures the effect of *G* on *X, r* measures the causal effect of *X* on *Y*, and *ε*_*X*_ and *ε*_*Y*_ 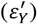 are independently and normally distributed random errors. Here, we used the term *r* for a causal effect of *X* on *Y* because it is equivalent to the correlation coefficient for two standardized phenotypes.

In the above model, the parameter *r* is the focus of MR analysis. In a typical two-sample MR analysis, regression coefficients of both *X* and *Y* on *G* are available from GWAS analyses, denoted by 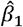 and 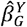. Then an unbiased estimator of *r* would be [23]

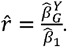

Below, we modeled the estimation problem using the genetic association z-score, which is defined as the regression coefficient divided by its standard error. Specifically,

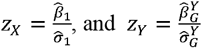

where 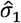 and 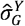 are standard errors of 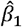 and 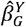.

The squared *z*-score statistic, termed *s*_*X*_ or *s*_*Y*_, is commonly used to test the genetic association between *G* and *X* or *Y*. When there is no association, the *s* statistic asymptotically follows a 1-df central chi-squared distribution. Accordingly, the *z*-score asymptotically follows a standard normal distribution. In contrast, when genetic association exists, the *s* statistic asymptotically follows a 1-df non-central chi-squared distribution characterized by a non-central parameter (NCP) *λ*, and the *z*-score asymptotically follows a normal distribution with variance one but with a non-zero mean parameter *μ* where *μ*^2^=*λ*, i.e.,

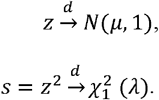

In our previous study, we proved that the NCP parameter *λ* is a function of sample size *N* and SNP effect size *h*^2^ [40]. Specifically,

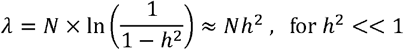

where ln() represents natural logarithm transformation, and *h*^2^ is the portion of phenotypic variance explained by the SNP, e.g., the SNP-specific heritability. Specifically, we had

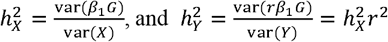

so that the mean parameters for *X* and *Y* have the following forms

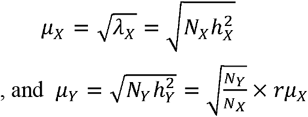

Clearly, the mean parameter *μ*_*Y*_ is completely determined by *μ*_*X*_ and their causal effect *r*.

In the above formula, the parameter *μ*_*X*_ is unknown and could be estimated from the GWAS summary statistics with the following formula

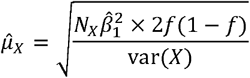

where *f* is minor allele frequency (MAF) of the IV. However, in practical released GWAS summary statistics *X* may not be standardized so that its variance var(*X*) is unknown. In this case, recall that

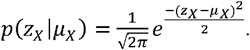

Therefore, we replaced *μ*_*X*_ by its maximum likelihood estimation, which is *z*_*X*_, and re-wrote the form of *μ*_*Y*_

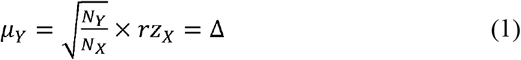

Given Δ, the probability of observing *z*_*Y*_ is simply the density of a normal distribution *N*(Δ, 1). Equivalently, if we let 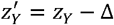, then 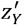 follows a standard normal distribution *N*(0, 1)

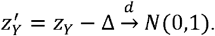

### Estimating *r* using z-scores

When multiple independent IVs are available, we built the estimation model within the maximum likelihood framework. Assumed that there are a total of *M* independent IVs. Let *Z*_*X*_ = (*Z*_*X1*_,*Z*_*X2*_,*Z*_*XM*_)’s and *Z*_*Y*_ = (*Z*_*Y1*_,*Z*_*Y2*_,*Z*_*YM*_)′ be corresponding z-score vectors.

We constructed the likelihood function of *r* as

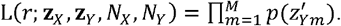

Maximizing the likelihood function L(*r*) yielded the maximum likelihood estimate of *r*, denoted by 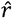, and the maximum likelihood, denoted by 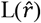.

The significance of 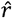 was evaluated using the LRT approach. Specifically, by restricting *r*=0, we obtained the null likelihood L_0_. We then constructed the likelihood ratio test statistic as follows

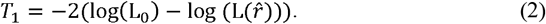

Under the null hypothesis of *r*=0, *T*_1_ approximately follows a 1-df central chi-squared distribution, which is used to judge the significance of 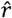.

### Modeling uncorrelated and correlated horizontal pleiotropy effects

We defined an IV to be valid if it has no horizontal pleiotropic effect. For a pleiotropic IV, the estimator 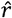 is not necessarily an unbiased estimator of *r* anymore. To see this, we re-wrote the phenotype model for *Y* as

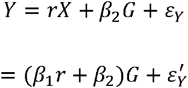

where *β*_2_ measures the pleiotropic effect of *G* on *Y* that is not mediated by *X*. In this model, 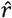 is an unbiased estimator of the following parameter instead

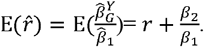

Depending on the direction of *β*_2_, 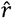 could overestimate or underestimate the true effect *r*.

In the presence of pleiotropy, the association of *G* with *Y* is a mixture of the *X*-mediated causal effect and the pleiotropic effect. Accordingly, the mean parameter *μ*_*Y*_ is a mixture of two components (**Supplemental Notes**)

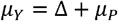

so that

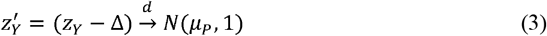

where 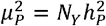 is a parameter defined by pleiotropy driven heritability

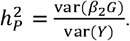

In the above formula, the mean parameter *μ*_*Y*_ is composed of two components: the first one is the causal effect, and the second one is the pleiotropic effect. The causal effect is fixed for not only valid IVs but also pleiotropic IVs. In contrast, the pleiotropic effect exists at pleiotropic IVs only and may vary depending on the strength of the IV-outcome effect, and was therefore modeled as a random effect. After removing the causal effect, the residual outcome z-score contains information about the pleiotropic effect only. Depending on whether it is correlated to *z*_*X*_ or not, the pleiotropic effect is further classified into two types. In the first type, the pleiotropic effect is directly on the outcome, so it is uncorrelated to *z*_*X*_. In this case, *μ*_*p*_ was modeled as a *z*_*X*_-independent normally distributed effect

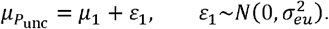

In the second type, the pleiotropic effect is influenced by some unmeasured confounder that is correlated to both the exposure and the outcome. *μ*_*p*_ in this type is correlated to *z*_*X*_ and the INSIDE assumption is violated [39]. Accordingly, it was modeled as a *z*_*X*_-related normally distributed effect

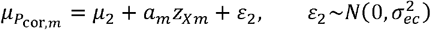

where *a*_*m*_ measures the correlation between 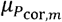 and *Z*_*Xm*_ at the *m*th IV. We added the subscript *m* in 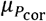 to emphasize that *a*_*m*_ is IV-specific whose magnitude depends on the magnitude of the pleiotropy effect at the *m*th IV. To account for the variation of *a*_*m*_, it was further modeled as a normally distributed random effect

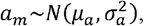

where *μ*_*a*_ and 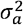 measure mean pleiotropic correlation and its variance, respectively.

Until now, we have built a hierarchical model towards the mean parameter of 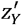 distribution. We aimed to obtain its marginal distribution. To integrate out the intermediate dummy variables, we introduced the following lemma,

*Lemma I, If a random variable X follows a normal distributio*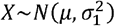, *and if its mean parameter *μ* is another random variable following a second normal distribution* 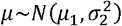, *then the marginal distribution of X is again a normal distribution of the form* 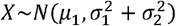 (**Supplemental Notes**).

Applying this lemma, we first integrated out *a*_*m*_ to obtain the marginal distribution of 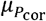

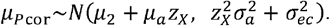

We then in turn integrated out *μ*_*punc*_ and *μ*_*pcar*_ to obtain the marginal distribution of 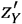

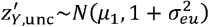

and

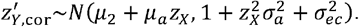

Taken together, for the three types of IVs (valid, uncorrelated pleiotropic and correlated pleiotropic), the residual z-score 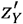 after removing the causal effect *r* has a type-specific distribution

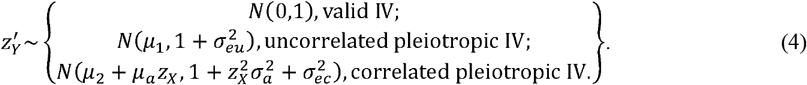

One key inference from the above distributions is that 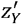 at different types has distinct variances. Specifically,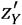 at valid IVs has an exact variance of one. 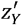 at pleiotropic IVs has a greater variance. This feature may make the three types of IVs distinguishable.

### Likelihood function under horizontal pleiotropy

We assumed that the proportions of valid IVs, uncorrelated pleiotropic IVs and correlated pleiotropic IVs are(1-*τ*), *τ* (1*-ρ*), and *τ ρ*, where *τ* and *ρ* ∈[0,1] measure the proportion of pleiotropic IVs and the relative proportion of correlated pleiotropic IVs, respectively. Our model contains nine parameters in total: 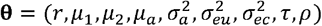. We constructed a likelihood function of **θ** as follows

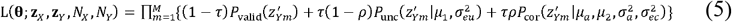

where the probability densities were defined by equation (4).

Maximizing L regarding **θ** yields the maximum likelihood estimate of **θ**, denoted by 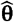, and the maximum likelihood, denoted by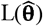. We maximized L using the function *nmkb* in the R package *dfoptim*, which implements a derivative-free Nelder–Mead algorithm for high-dimensional function.

### Test of causal effect

The causal effect is tested against the null hypothesis of r=0 by means of the LRT approach. Specifically, by restricting *r*=0 in (5), we constructed the null likelihood function, enoted by L_0_, which contains eight parameters. Maximizing L_0_ over its sample space yields the maximum likelihood under the null hypothesis, denoted by 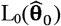. The LRT statistic was then constructed as follows

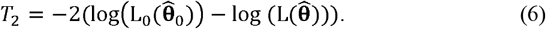

Under the null hypothesis of *r*=0, *T*_2_ approximately follows a 1-df central chi-squared distribution, which was used to declare the significance of 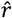.

### Test of pleiotropic effects

The existence of pleiotropic effects was tested against the null hypothesis of *τ*=0 using the same LRT approach. Under the null hypothesis, the likelihood function contains only one free parameter *r*. Therefore, we used a 8-df central chi-squared distribution to declare the significance of the estimate 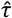.

In implementation, we will first test pleiotropic effects at the significance level *P*<0.05. If there is evidence of pleiotropic effects, then we will use *T*_2_ to test the causal effect; otherwise, we will use *T*_1_ instead.

### Unstandardized phenotypes

Since 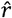 is estimated based on z-scores, its test significance remains unchanged regardless of whether exposure or outcome is standardized. However, its magnitude may differ from the original effect size when unstandardized exposure or outcome is analyzed. The ratio of 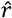 to the original effect size is a constant *C* that is determined by the unit of exposure and outcome under analysis. In certain scenarios, it is desirable to obtain the original effect size, such as to recover the odds ratio for case-control data types.

To recover the original effect size, we estimated *C* by anchoring our test to a comparable alternative test. As demonstrated in the Results section, our proposed test statistic *T*_1_, which assumes no pleiotropic effects, yields an effect size approximately equal to the IVW test, which also assumes no pleiotropic effects. Consequently, we conducted both the *T*_1_ test and the IVW test on the same dataset and estimated *C* by calculating the ratio of their effect sizes.

### Simulation studies

We performed a series of simulation studies to evaluate the performance of the proposed method. We simulated one continuous exposure *X*, one continuous outcome *Y* and one continuous confounder *U*. We simulated a set of bi-allelic SNP as IVs for the exposure. We studied a two-sample context so that the exposure and outcome summary statistics were generated from two separate samples. Parameter settings were as follows:

1. The number of IVs was set to be *M*=200.
2. Both exposure and outcome sample sizes were set to be *N*_*X*_*=N*_*Y*_=200,000.
3. The causal effect of *X* on *Y* was set to be *r*=0 (null), 0.05 (positive effect), or -0.05 (negative effect). In the latter two cases, *X* explained *r*^2^=0.25% of *Y’*s variance.
4. Confounder effect. In the case of confounding effect, the effect of the confounder *U* was set to explain 20% of phenotypic variance in both *X* and *Y*. The effect directions of *U* on both *X* and *Y* were set to be the same.
5. Proportion of pleiotropic IVs to the outcome (*τ*). The proportion of pleiotropic IVs *τ* varied from 10% to 60% at an increment of 10%. The default direction of the pleiotropic effects was assumed to be positively dominated, in which 70%: 30% of pleiotropic IVs were simulated to have positive: negative pleiotropic effects. In the case of balanced pleiotropic effects, 50%: 50% of pleiotropic IVs were simulated to have positive: negative pleiotropic effects.
6. Relative proportion of correlated IVs (*ρ*). From the pleiotropic IVs simulated in step 5), a fraction of *ρ* = 0.5 IVs were simulated to also have a pleiotropic effect on *U*. For ease of presentation, the direction of the pleiotropic effects on *U* was set to be the same as that on *Y*.
7. IV effects on exposure. We assumed a non-infinitesimal genetic architecture for IV-exposure effects. A proportion of 10% of total IVs were simulated to have a large effect each explaining 0.2% of *X*’s variance. The portion of explained variance for each of the remaining 90% IVs was drawn from an exponential distribution with a mean 1.0 10^−4^, a level comparable to the majority of GWAS findings. The 200 IVs explained approximately a total of 5.8% of *X*’s variance.
8. IV pleiotropic effects on outcome and confounder. The portion of the variance in both *Y* and *U* explained by each pleiotropic IV was drawn from an exponential distribution with a mean 1.0 × 10^−4^.

Each scenario was simulated with 1,000 iterations. At each simulation iteration, the MAF of each IV was drawn from a uniform distribution *uni*(0.05, 0.5), and the IV genotypes at both exposure and outcome samples were simulated using PLINK [59] assuming the Hardy-Weinberger equilibrium. After the phenotypes were simulated, PLINK was invoked to test genetic association in both the exposure and the outcome samples.

### Comparison with existing methods

We evaluated and compared the performance of the proposed method with 14 existing two-sample MR methods, including IVW [23], MREGGER [22], weighted-median (W-median) [21], weighted-mode (W-mode) [20], MRPRESSO [19], the contamination mixed model (CMix) [16], MRMix [18], MRAID [11], MRcML [12], GRAPPLE [13], MRLASSO [17], MRRAPS [14], MRROBUST [17], and CAUSE [15]. Comparison criteria included type-I error rate, power rate and estimated effect size. The statistical significance was declared at the nominal level *α*=0.05. In power rate estimation, only effects in the same direction as the true effect were considered as potentially successful hits. Effects in the opposite direction were not considered regardless of their statistical significance.

Because various methods may possess different type-I error rates, comparing raw power rates may be unfair. To ensure a fair comparison of power rates, the raw power rate was corrected by calibrating it with the type-I error rate estimated at the corresponding null condition. This correction was only for comparison purpose. It allows all methods to operate under the assumption of possessing a correct type-I error rate of 0.05

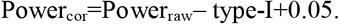

CAUSE requires a substantial number of background SNPs spanning the genome to estimate its model parameters. To meet this requirement, we generated an additional random set of 100,000 independent SNPs that are associated with neither the exposure nor the outcome. This combination of 100,000 background SNPs and the IVs was used for estimating nuisance parameters (step 1), while only the IVs were used for causal inference (step 2). For all other methods, only the IVs were used for causal inference.

Due to their high computational demands, CAUSE and MRPRESSO were assessed across 200 iterations.

### Real data application

Lean body mass is an important physiological index. Low ALM, coupled with diminished muscle strength and reduced physical performance, serves as a defining criterion for the onset of sarcopenia [60], which is a critical condition that can significantly impair function, lead to physical disability, and is a major modifiable risk factor for frailty in older adults [61, 62]. Lipids are another cluster of metabolites related to energy balance [63, 64]. Disorders of lipid metabolism can co-occur with the loss of skeletal muscle mass [65, 66]. However, the mutual relationship between ALM and lipid traits has not been well-studied. Uncovering their causal relationships is thus needed to facilitate the prediction and intervention of sarcopenia.

As a real application, we conducted a comprehensive bidirectional MR study between ALM and four lipid traits, including HDL-C, LDL-C, TC, and TG using two data sources: the UKB internal cohort and the summary statistics released by GLGC. The study (project number 41542) was covered by general ethical approval for the UKB study, and was approved by the Northwest Centre for Research Ethics Committee (11/NW/0382). All participants provided informed consent.

The study design is displayed in **Supplemental Figure 7**. In brief, we performed both the UKB-internal analysis and the UKB-GLGC joint analysis. In the UKB-internal analysis, we randomly divided the entire UKB cohort into two independent sub-samples (UKB_S1 and UKB_S2). For each exposure-outcome pair (e.g., ALM-TC), one sub-sample served as the exposure sample, while the other served as the outcome sample. This process was repeated by reversing the roles of the two sub-samples so that two independent sets of summary statistics were generated [41]. In the UKB-GLGC analysis, the whole UKB cohort was used to generate GWAS summary statistics for ALM, while the GLGC data (excluding UKB participants) provided summary statistics for the lipid traits. In total, for each exposure-outcome pair, we generated three datasets that are mutually independent in exposure and/or outcome, so that the results from them could cross-validate each other.

To evaluate the influence of sample overlap, we also applied a MR analysis in which both the exposure and the outcome summary statistics were derived from the whole UKB cohort, so that both samples completely overlapped.

The details of the analysis are described in **Supplemental Notes**. In brief, the GWAS of UKB samples was performed with BOLT-LMM [67]. Genome-wide significant (p<5.0 × 10^−8^) SNPs were selected from the exposure sample, followed by clumping (LD r^2^=0.01 and window size=500 kb) to select eligible IVs using *TwoSampleMR* R package. In the UKB-GLGC joint analysis, GWAS of ALM was conducted in the whole UKB cohort, while the GWAS summary statistics of lipid traits were derived from the released GLGC results including no UKB participants. In the UKB-overlapping analysis, the GWAS summary statistics of both ALM and lipid traits were derived from the whole UKB cohort. The QC and MR analysis procedure are the same across various analyses. However, in analyses using UKB samples only (UKB-internal and UKB-overlapping), palindromic IVs were not excluded, while in the UKB-GLGC joint analysis, palindromic IVs were excluded to avoid strand orientation error.

### Mouse-model experiments

The experimental procedures and treatments conducted in this study were ethically reviewed and approved by the Animal Care Ethical Committee of Soochow University, Suzhou, China, ensuring compliance with animal welfare guidelines and regulations.

Male C57BL/6 mice, aged 8 weeks and weighing 20 ± 5 g, were procured from Shanghai Lingchang Biotechnology Co., LTD. These mice were housed in a controlled environment with a temperature of 23 ± 1 °C and a 12:12-hour light-dark cycle (lights on from 07:00 to 19:00), and they had free access to food and water throughout the experiment. Following a 1-week acclimatization period, the mice were randomly assigned to two groups: the control group (*N*=12) and the TC group (*N*=12). Standard feed was obtained from Double Lion Experimental Animal Feed Technology Co., LTD, Suzhou. The control group received a standard diet consisting of 71% normal diet food, 20% protein, 4% fat, and 5% fiber. In contrast, the TC group received a high-cholesterol diet comprising 68.3% normal diet food, 1.3% cholesterol, 18.4% lard, and 12% protein. This dietary intervention continued for a duration of 8 weeks.

Body composition analysis, including measurements of total body mass, fat mass, lean mass, and fluid content, was conducted on live animals without the use of anesthesia. This analysis was performed using small animal MRI equipment (Minispec LF50 body composition analyzer, Bruker, Billerica, MA, USA). To conduct the measurements, each mouse was placed in a specially designed plastic holder tailored for mice, without the need for sedation or anesthesia. Subsequently, the holder containing the mouse was positioned within the measuring space of the MRI system. To ensure the accuracy of the results, measures were taken to prevent the mice from moving within the holder during the scanning process. Each scan lasted approximately 2 minutes, during which the MRI equipment captured detailed data on the body composition of the mice.

Blood biochemistry analysis was conducted using a Hitachi 7100 clinical chemistry analyzer in accordance with the manufacturer’s guidelines. Approximately 500 *μ*L of plasma was collected from each mouse and transferred to a gel tube containing lithium heparin. Subsequently, the plasma samples were centrifuged at 5000 rpm using a refrigerated centrifuge set at 4°C for 15 minutes to obtain 160–200 *μ*L of serum. In cases where the volume of serum obtained was insufficient for analysis, it was diluted with deionized water at a ratio of 1:2 to ensure proper loading for analysis. This process ensured accurate and reliable blood biochemistry measurements for each sample.

For each trait, the difference of measurement between the endpoint and the baseline was analyzed. The comparison was made between the control group and the intervention group using the Wilcoxon rank test in R package.

## Data Availability

Access to UK Biobank data can be obtained by application to UK Biobank (https://www.ukbiobank.ac.uk/).

## Code availability

All analyses have been performed using publicly available software or custom codes. PLINK (v1.90b6.5, https://www.cog-genomics.org/plink/) and BOLT-LMM (v2.3.2, https://alkesgroup.broadinstitute.org/BOLT-LMM/) were used to perform association analysis in the simulated data and in the UK Biobank data, respectively. The MR analyses were performed using R (v.4.3.2, https://cran.r-project.org/). The R package TwoSampleMR (v0.5.7, https://github.com/MRCIEU/TwoSampleMR) was used to implement the IVW, MREGGER, W-median, and W-mode methods. MRAID (https://github.com/yuanzhongshang/MRAID), CAUSE (v1.2.0, https://github.com/jean997/cause), MRcML (https://github.com/xue-hr/MRcML/), GRAPPLE (https://github.com/jingshuw/GRAPPLE), MRMix (https://github.com/gqi/MRMix), mr.raps (https://github.com/qingyuanzhao/mr.raps), MR-PRESSO (https://github.com/rondolab/MR-PRESSO) were used to implement respective methods. MendelianRandomization (v0.9.0, https://cran.r-project.org/web/packages/MendelianRandomization/index.html) was used to implement the CMix, MRLASSO and MRROBUST methods [68]. The R package dfoptim (v2023.1.0, https://cran.r-project.org/web/packages/dfoptim/index.html) was used to optimize high-dimensional likelihood function.

## Acknowledgments

This research was conducted using the UK Biobank resource under application number 41542. Special thank was given to an anonymous reviewer whose constructive comments advised us to model correlated pleiotropic effects. This study was partially supported by the funding from National Natural Science Foundation of China (32170670 to YFP, 81902181 to JJN), the Project of MOE Key Laboratory of Geriatric Diseases and Immunology (JYN202401 to HGR), and Suzhou Basic Research Program (Medical Application Basic Research) (SKY2023147 to JJN).

