## supplemental notes for "A new robust and accurate two-sample Mendelian randomization method with a large number of genetic variants"

1. **Decomposition of mean parameter** $\boldsymbol{\mu}_{\boldsymbol{Y}}$ **into two components**

In the presence of horizontal pleiotropy, the phenotype model of *Y* is expressed as

$$Y=rX+\beta_{2}G+\varepsilon_{Y}=(\beta_{1}r+\beta_{2})G+\varepsilon_{Y}^{'}$$

, so that the mean parameter $\boldsymbol{\mu}_{\boldsymbol{Y}}$ for *z_Y_* is

$\mu_{Y}^{2}=N_{Y}h_{Y}^{2}=N_{Y}\frac{\left( \beta_{1}r+\beta_{2} \right)^{2}var(G)}{var(Y)}$.

Without loss of generality, we assumed that both $r>0$ and $\beta_{2}>0$, so that

$$\mu_{Y}=\sqrt{\frac{N_{Y}\beta_{1}^{2}var(G)}{var(Y)}}r+\sqrt{\frac{N_{Y}\beta_{2}^{2}var(G)}{var(Y)}}$$

$=\sqrt{\frac{N_{Y}}{N_{X}}}\sqrt{\frac{N_{X}\beta_{1}^{2}var(G)}{var(Y)}}r+\sqrt{\frac{N_{Y}\beta_{2}^{2}var(G)}{var(Y)}}$.

By definition, var(*X*)=var(*Y*)=1, so that

$$\mu_{Y}=\sqrt{\frac{N_{Y}}{N_{X}}}\sqrt{\frac{N_{X}\beta_{1}^{2}var(G)}{var(X)}}r+\sqrt{\frac{N_{Y}\beta_{2}^{2}var(G)}{var(Y)}}$$

$=\sqrt{\frac{N_{Y}}{N_{X}}}\mu_{X}r+\mu_{P}$.

**2.	Proof of Lemma I**

*Lemma I, If a random variable X follows a normal distribution* $X\sim N\left( \mu,\sigma_{1}^{2} \right)$*, and if its mean parameter* $\mu$ *is another random variable following a second normal distribution* $\mu\sim N\left( \mu_{1},\sigma_{2}^{2} \right)$*, then the marginal distribution of X is again a normal distribution of the form* $X\sim N\left( \mu_{1},\sigma_{1}^{2}+\sigma_{2}^{2} \right)$

**Proof**: Recall that $p\left( X | \mu_{1},\sigma_{1}^{2},\sigma_{2}^{2} \right)=\int_{-\infty}^{\infty} p\left( X | \mu,\sigma_{1}^{2} \right)p\left( \mu| \mu_{1},\sigma_{2}^{2} \right)d\mu$, we derive the marginal distribution of *X* by integrating out $\mu$

$$p\left( X | \mu_{1},\sigma_{1}^{2},\sigma_{2}^{2} \right)=\int_{-\infty}^{\infty} \frac{1}{\sqrt{2\pi\sigma_{1}^{2}}}exp\{-\frac{{(X-\mu)}^{2}}{2\sigma_{1}^{2}}\}\cdot\frac{1}{\sqrt{2\pi\sigma_{2}^{2}}}exp\{-\frac{{(\mu-\mu_{1})}^{2}}{2\sigma_{2}^{2}}\}d\mu$$

$$=\int_{-\infty}^{\infty} \frac{1}{2\pi\sqrt{\sigma_{1}^{2}\sigma_{2}^{2}}}exp\{-\frac{{(X-\mu)}^{2}}{2\sigma_{1}^{2}}-\frac{{(\mu-\mu_{1})}^{2}}{2\sigma_{2}^{2}}\}d\mu$$

$$=\int_{-\infty}^{\infty} \frac{1}{2\pi\sqrt{\sigma_{1}^{2}\sigma_{2}^{2}}}exp\{-\frac{{\sigma_{2}^{2}(X-\mu)}^{2}+{\sigma_{1}^{2}(\mu-\mu_{1})}^{2}}{2\sigma_{1}^{2}\sigma_{2}^{2}}\}d\mu$$

$$=\int_{-\infty}^{\infty} \frac{1}{2\pi\sqrt{\sigma_{1}^{2}\sigma_{2}^{2}}}exp\{-\frac{{{(\sigma}_{1}^{2}+\sigma}_{2}^{2})\mu^{2}-2\left( X\sigma_{2}^{2}+\mu_{1}\sigma_{1}^{2} \right)\mu+{\sigma_{2}^{2}X}^{2}+\sigma_{1}^{2}\mu_{1}^{2}}{2\sigma_{1}^{2}\sigma_{2}^{2}}\}d\mu$$

$$=\int_{-\infty}^{\infty} \frac{1}{2\pi\sqrt{\sigma_{1}^{2}\sigma_{2}^{2}}}exp\{-\frac{{(\mu-\frac{X\sigma_{2}^{2}+\mu_{1}\sigma_{1}^{2}}{{\sigma_{1}^{2}+\sigma}_{2}^{2}})}^{2}+\frac{{\sigma_{2}^{2}X}^{2}+\sigma_{1}^{2}\mu_{1}^{2}}{{\sigma_{1}^{2}+\sigma}_{2}^{2}}-{(\frac{X\sigma_{2}^{2}+\mu_{1}\sigma_{1}^{2}}{{\sigma_{1}^{2}+\sigma}_{2}^{2}})}^{2}}{\frac{2\sigma_{1}^{2}\sigma_{2}^{2}}{{\sigma_{1}^{2}+\sigma}_{2}^{2}}}\}d\mu$$

$$=\int_{-\infty}^{\infty} \frac{1}{2\pi\sqrt{\sigma_{1}^{2}\sigma_{2}^{2}}}\exp\left\{ -\frac{\left( \mu-\frac{X\sigma_{2}^{2}+\mu_{1}\sigma_{1}^{2}}{{\sigma_{1}^{2}+\sigma}_{2}^{2}} \right)^{2}}{\frac{2\sigma_{1}^{2}\sigma_{2}^{2}}{{\sigma_{1}^{2}+\sigma}_{2}^{2}}} \right\}d\mu\cdot exp\{-\frac{({\sigma_{2}^{2}X}^{2}+\sigma_{1}^{2}\mu_{1}^{2})({\sigma_{1}^{2}+\sigma}_{2}^{2})-{(X\sigma_{2}^{2}+\mu_{1}\sigma_{1}^{2})}^{2}}{2\sigma_{1}^{2}\sigma_{2}^{2}({\sigma_{1}^{2}+\sigma}_{2}^{2})}\}$$

$$=\frac{1}{2\pi\sqrt{\sigma_{1}^{2}\sigma_{2}^{2}}}\cdot\sqrt{2\pi\left( \frac{\sigma_{1}^{2}\sigma_{2}^{2}}{{\sigma_{1}^{2}+\sigma}_{2}^{2}} \right)}\cdot exp\{-\frac{\left( X-\mu_{1} \right)^{2}}{2\left( {\sigma_{1}^{2}+\sigma}_{2}^{2} \right)}\}$$

$=\frac{1}{\sqrt{2\pi({\sigma_{1}^{2}+\sigma}_{2}^{2})}}\cdot exp\{-\frac{\left( X-\mu_{1} \right)^{2}}{2\left( {\sigma_{1}^{2}+\sigma}_{2}^{2} \right)}\}$,

which is the probability density of the normal distribution $N\left( \mu_{1},\sigma_{1}^{2}+\sigma_{2}^{2} \right)$.

**3.	Analysis of the UKB cohort data**

Phenotype measurement of appendicular lean mass (ALM) was detailed elsewhere [1]. In brief, body composition was measured by the bioelectricalimpedance analysis (BIA)approach.ALM was quantified by the sum of fat-free mass at the arms (data fields 23121 and23125) and legs (data fields 23113 and 23117).Lipid traits were measured from the serum sample in the UKB central laboratory.

As strict quality control procedure, we only analyzed self-reported white participants (data field 21000). Participants who had a self-reported sex inconsistent with the genetic sex derived from sex-related genotype data, whose sex chromosome was aneuploid, who had unusually high heterozygosity, who had high missing genotype rates or who withdrew their consent were removed.

Phenotype outliers were examined in the two sex groups separately. In each sex, phenotypic outliers were monitored by the inter-quartile range (IQR) approach, and the outliers were excluded. The two groups after excluding outliers were combined. Covariates, including age, sex, the top 10 principal components (PCs) and assessment center (23 levels), were used to adjust phenotypes. The residuals after adjustment were transformed into the inverse quantiles of a standard normal distribution to ensure normality, which were used for all subsequent analyses.

Genome-wide genotypes were available for all participants at 784,256 genotyped autosome markers, and were imputed into UK10K haplotype, 1000 Genomes project phase 3 and Haplotype Reference Consortium (HRC) reference panels. A total of ~92 million variants were generated by imputation. We excluded variants with minor allele frequency (MAF)<1% and with imputation r^2^<0.3.

The entire UKB sample was randomly divided into two sub-samples, UKB_S1 and UKB_S2. Genome-wide association study was conducted in the entire sample and in the two sub-samples, respectively. For each phenotype, we used BOLT-LMM to perform linear mixed model (LMM) analysis [2]. As the LMM analysis can adjust for population structure and relatedness, we did not include principal components (PCs) of ancestry as covariates in the LMM analysis.

Genome-wide significant SNPs (*P*<5$\times$10^-8^) were selected, followed by clumping using a window size of 500 kb region and a *r*^2^ threshold of 0.01 to retain independent IVs. Palindromic SNPs were allowed in the UKB-internal analysis. The collective IV-attributable exposure variance was estimated using the following equation

$R^{2}=\sum\hat{\beta}^{2}\times2f(1-f)$,

where $\hat{\beta}$ and *f* are association coefficient and minor allele frequency. The *F*-statistic was estimated using the following equation

$$F=\frac{(N-k-1)\times R^{2}}{k\times(1-R^{2})}$$

, where *N* and k are sample size and the number of IVs, respectively.

4.	**The R code implementing the likelihood function of MRZ**

dat<-read.table('mr_file.txt',header=T,sep="\t")

library(dfoptim)

b.exp = dat$b.exp

b.out = dat$b.out

se.exp = dat$se.exp

se.out = dat$se.out

zx<-b.exp/se.exp

zy<-b.out/se.out

ny=dat$N.out

nx=dat$N.exp

sign = sign(zx)

zx = zx*sign

zy = zy*sign

density_valid<-function(r,zx,zy,nx,ny){

zy1=zy-sqrt(ny/nx)*zx*r

return(max(dnorm(38),dnorm(zy1,mean=0,sd=1)))

}

density_uncorrelated_pleiotropy<-function(r,zx,zy,nx,ny,mu,var){

zy1=zy-sqrt(ny/nx)*zx*r-mu

return(max(dnorm(38),dnorm(zy1,mean=0,sd=sqrt(var))))

}

density_correlated_pleiotropy<-function(r,a,zx,zy,nx,ny,mu,var){

zy1=zy-sqrt(ny/nx)*r*zx-sqrt(ny/nx)*a*zx-mu

return (max(dnorm(38),dnorm(zy1,mean=0,sd=sqrt(var))))

}

likelihood_full<-function(par,zx,zy,nx,ny){

mu1=par[1]

mu2=par[2]

veu=par[3]

vec=par[4]

va=par[5]

tao =par[6]

rho =par[7]

a=par[8]

r=par[9]

sum =0

for(i in 1:length(zy)){

z_x=zx[i]

z_y=zy[i]

n_x=nx[i]

n_y=ny[i]

density_valid = density_valid(r=r,zx=z_x,zy=z_y,nx=n_x,ny=n_y)

density_uncorrelated = density_uncorrelated_pleiotropy(r=r,zx=z_x,zy=z_y,nx=n_x,ny=n_y,mu=mu1,var=1+veu)

density_correlated = density_correlated_pleiotropy(r=r,a=a,zx=z_x,zy=z_y,nx=n_x,ny=n_y,mu=mu2,var=1+vec+va*z_x*z_x)

density = density_valid*(1-tao)+density_uncorrelated*tao*rho+density_correlated*tao*(1-rho)

sum = sum-log(density)

}

return(sum)

}

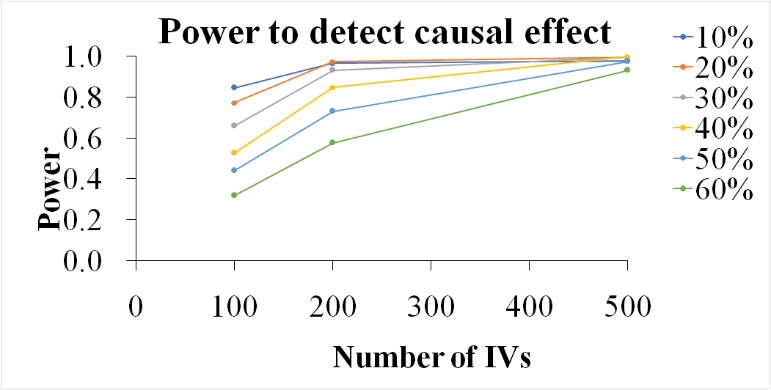

**Supplemental Figure 1. Power of MRZ with various numbers of IVs.**

The number of IVs varied between 100, 200, and 500. The sample sizes for both exposure and outcome samples were 2e5. The proportion of pleiotropic IVs ranged from 10%-60%. A positively dominated pleiotropic setting was simulated, in which 70% pleiotropic IVs were simulated to have positive pleiotropic effects while the remaining 30% were simulated to have negative pleiotropic effects. Half pleiotropic IVs were simulated to be correlated with the corresponding IV-exposure effects. The causal effect *r* was set to be 0.05 (positive effect). The statistical significance was declared at $\alpha=0.05$level.

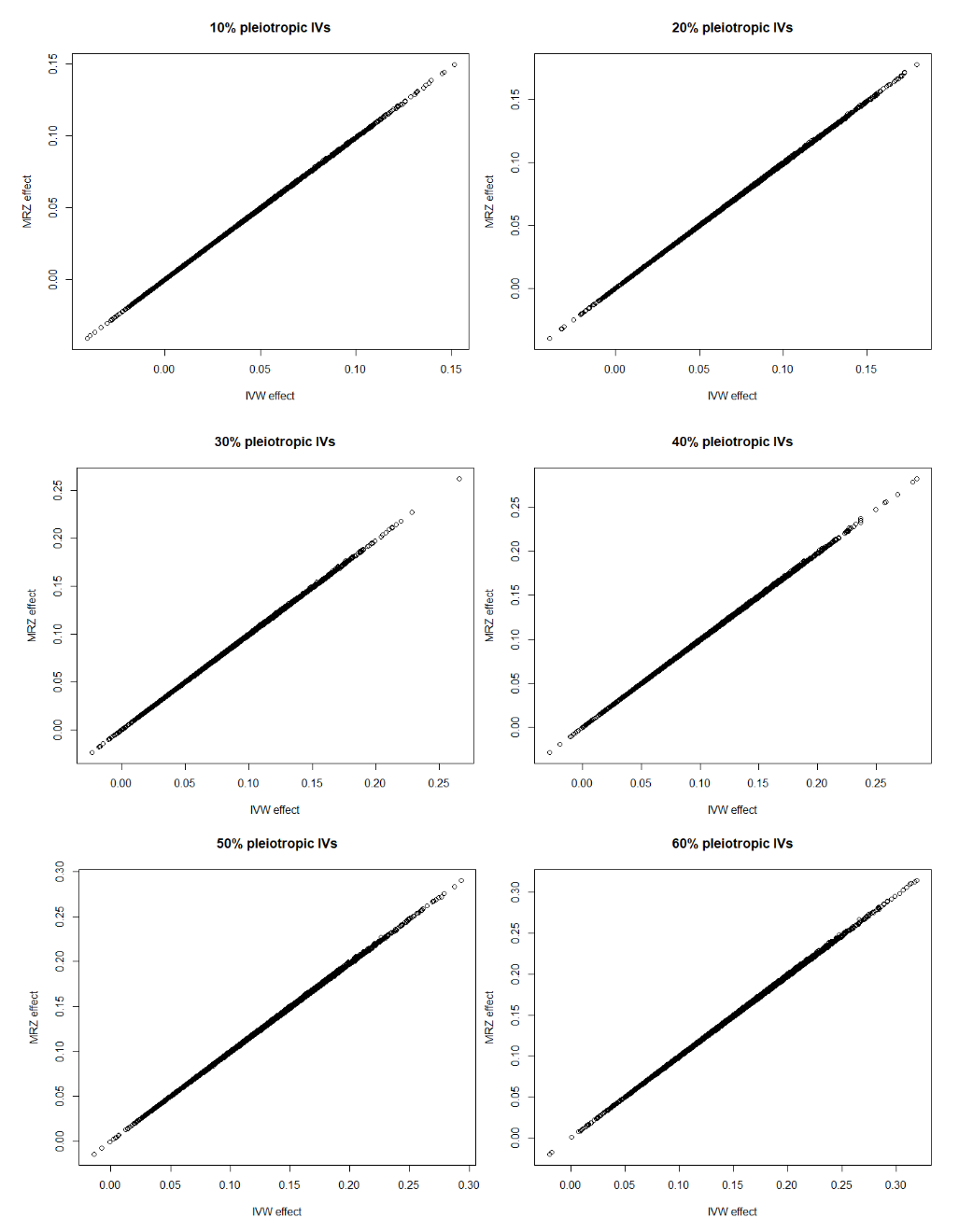

**Supplemental Figure 2. The concordance of effect size estimated by the proposed method and the IVW test.**

The proportion of pleiotropic IVs ranged from 10%-60%. Both causal and null simulations were simulated, whose results were summarized together for a particular proportion of pleiotropic IVs.

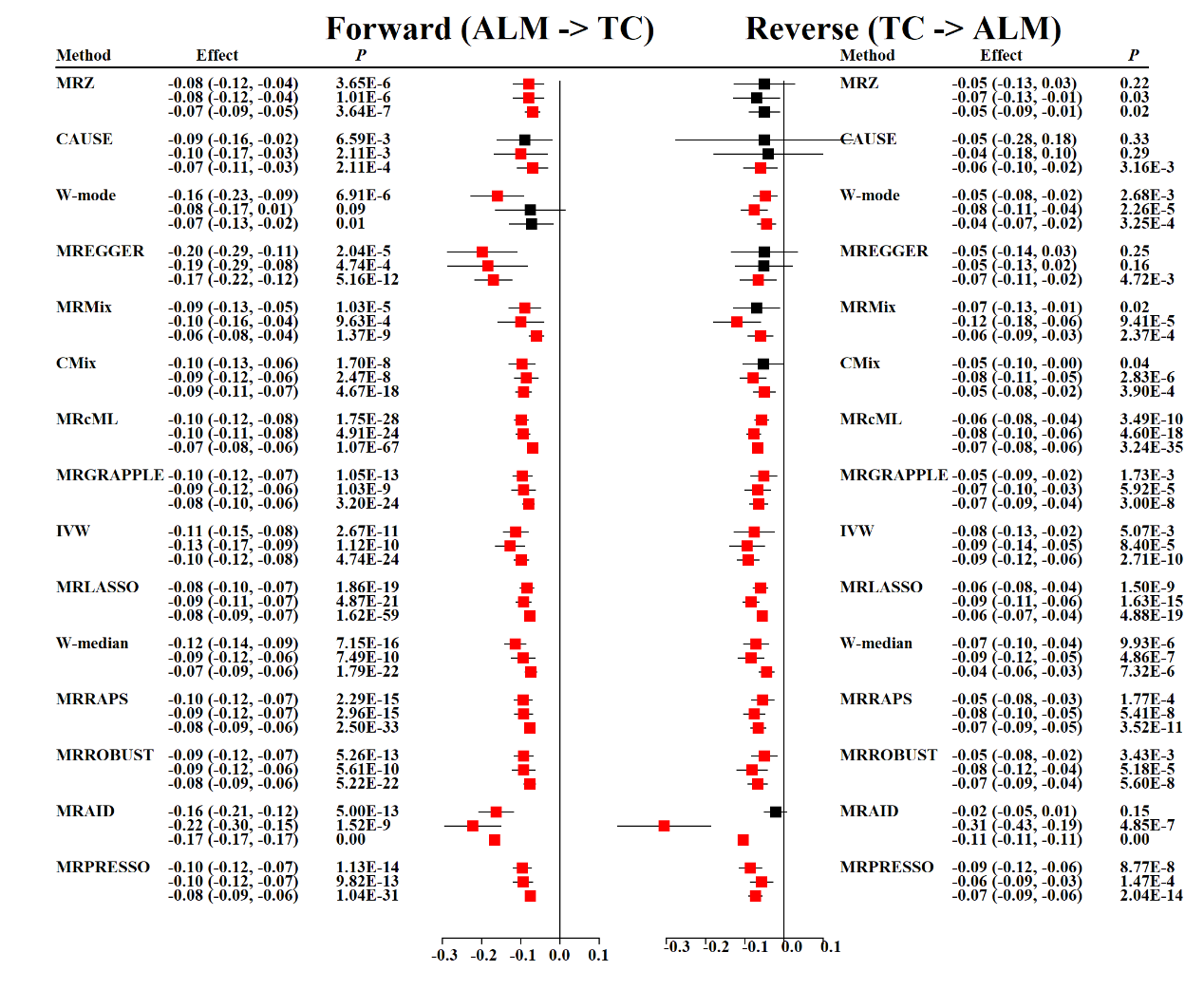

**Supplemental Figure 3. Bidirectional MR analysis results for ALM and TC.**

The entire UKB cohort was randomly divided into two independent sub-samples (UKB_S1 and UKB_S2). For the UKB-internal analysis, the two sub-samples were used as exposure and outcome samples, respectively. For the UKB-GLGC joint analysis, the entire UKB sample was used for ALM, while GLGC summary statistics were used for lipid traits. For each method, there are three datasets, namely UKB_S1/UKB_S2, UKB_S2/UKB_S1, and UKB/GLGC. Both forward (left) and reverse (right) MR analyses were performed. The significance threshold was set at 6.25$\times$10^-3^ (0.05/8). Significant associations were marked in red.

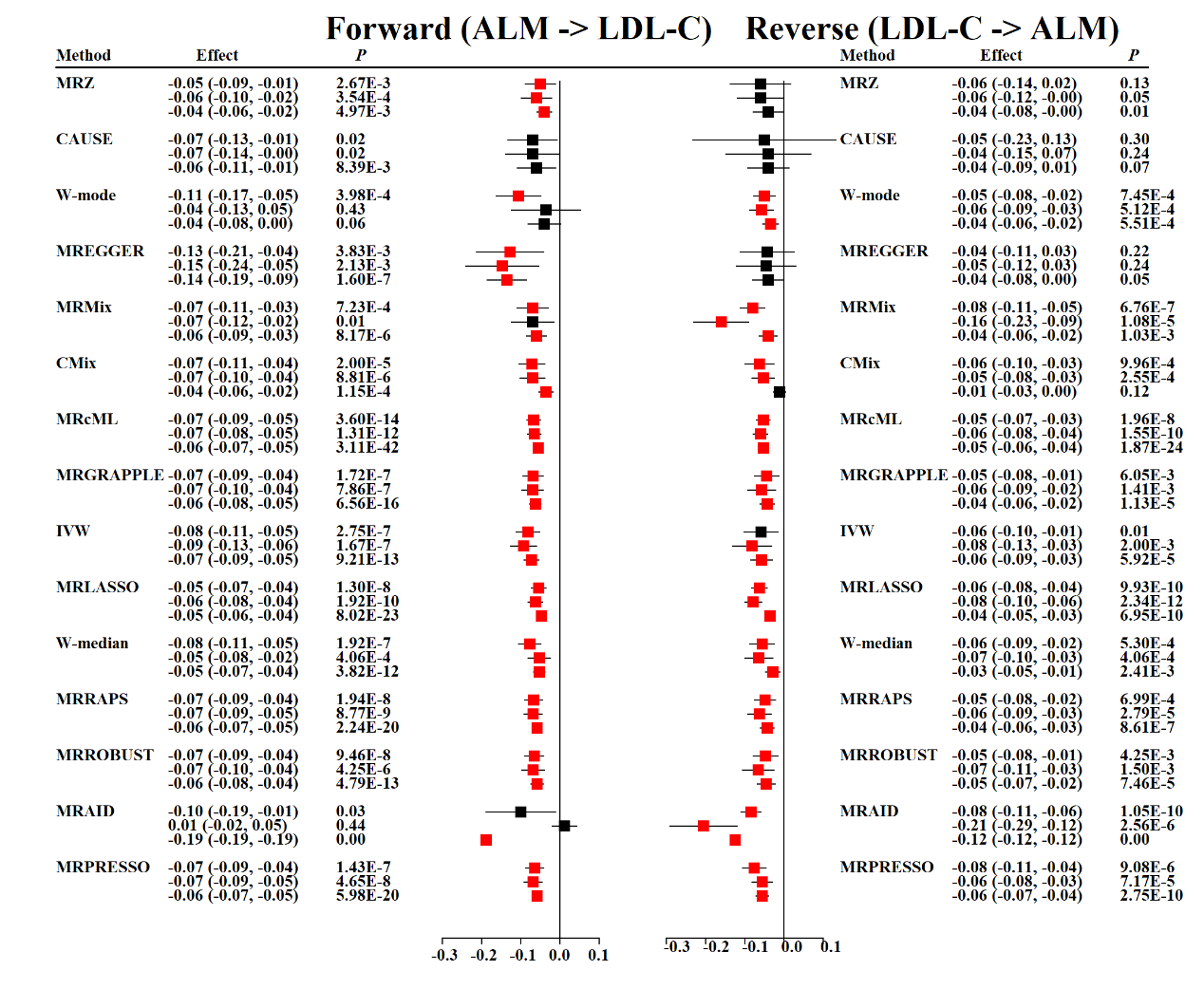

**Supplemental Figure 4. Bidirectional MR analysis results for ALM and LDL-C.**

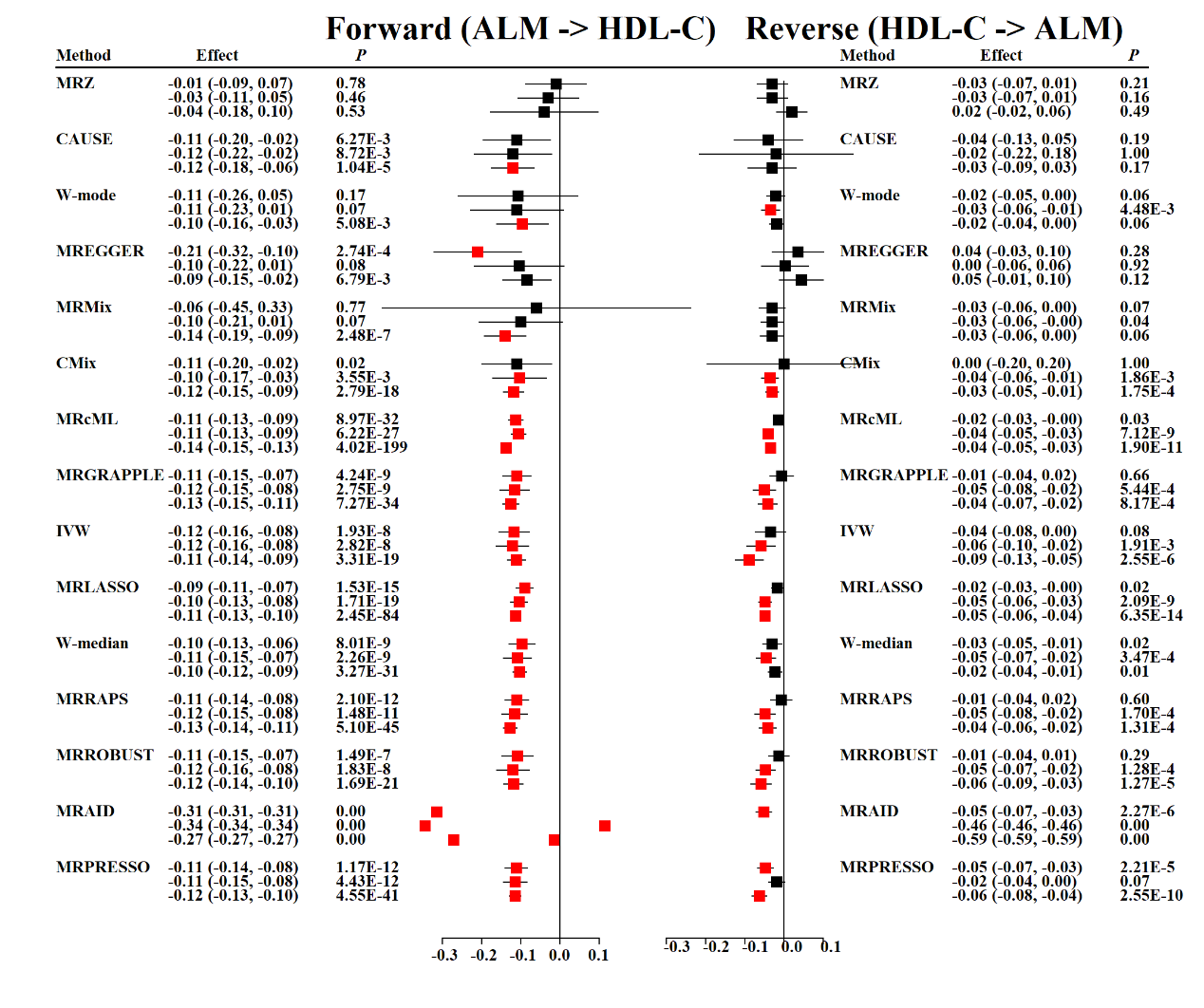

**Supplemental Figure 5. Bidirectional MR analysis results for ALM and HDL-C.**

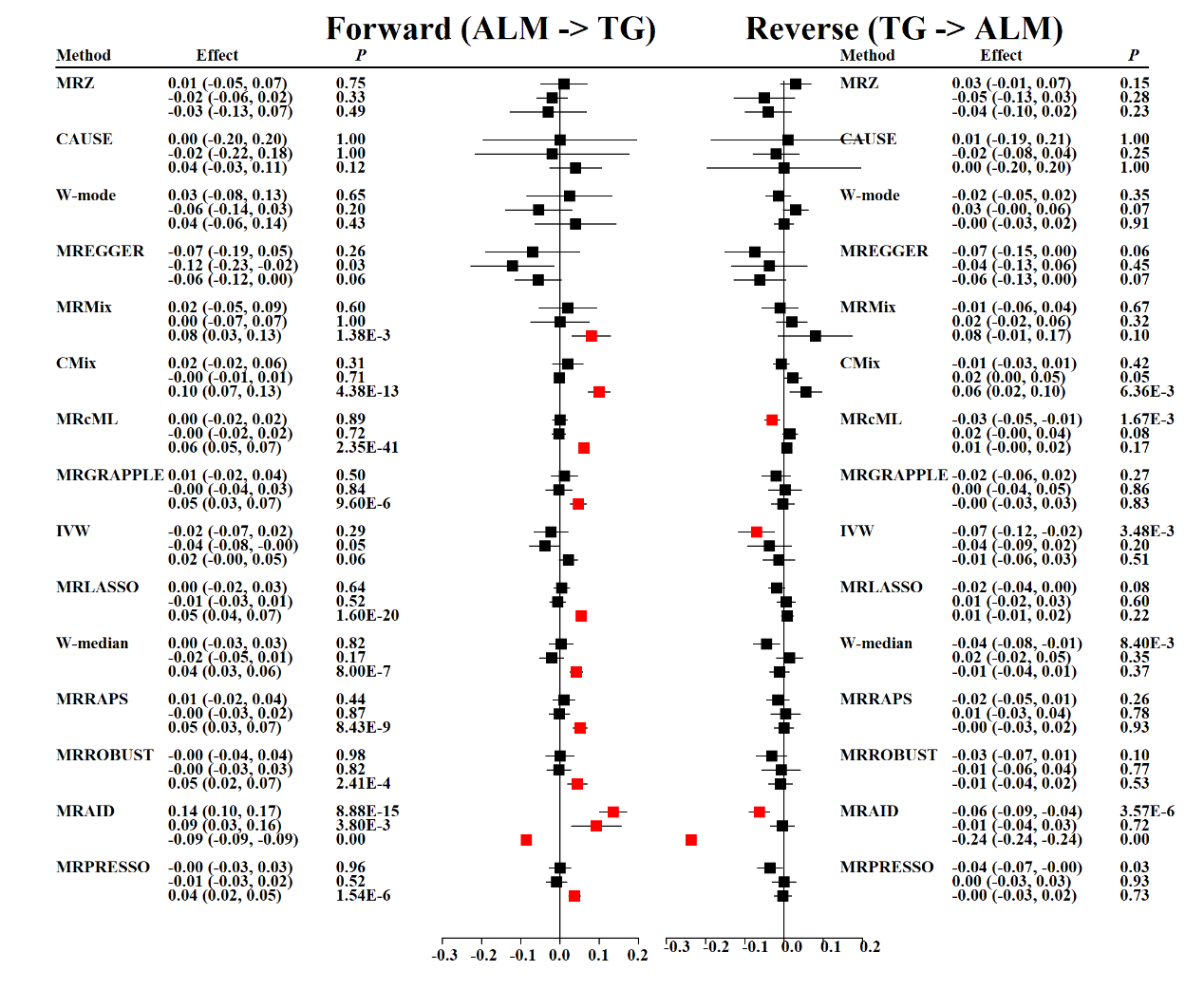

**Supplemental Figure 6. Bidirectional MR analysis results for ALM and TG.**

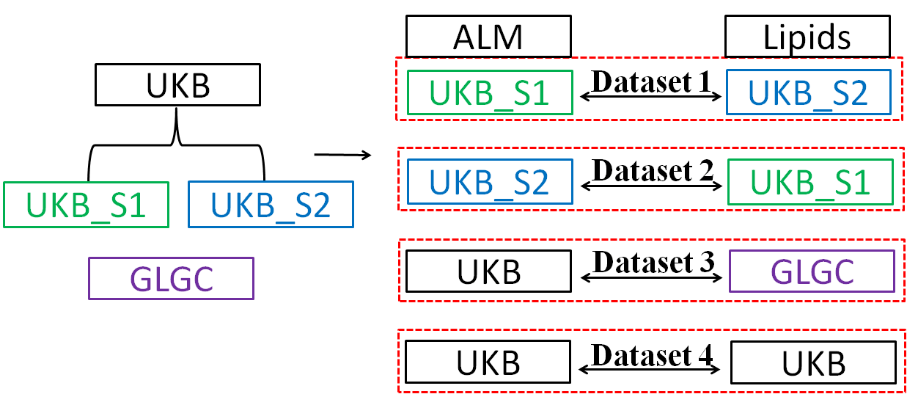

**Supplemental Figure 7. Study design for the real data MR analysis.**

The entire UKB cohort was randomly divided into two independent sub-samples (UKB_S1 and UKB_S2). In dataset 1, UKB_S1 served as ALM sample, while UKB_S2 served as lipid traits sample. In dataset 2, UKB_S2 served as ALM sample, while UKB_S1 served as lipid traits sample. In dataset 3, the entire UKB cohort served as ALM sample, while the GLGC summary statistics served as lipid traits sample. In each dataset, both forward and reverse MR analyses were performed.
